# Non-Invasive Brain Stimulation for Core Symptoms of Chronic Primary Pain: A Systematic Review and Meta-Analysis of randomized controlled trials

**DOI:** 10.1101/2025.04.17.25325770

**Authors:** Alessandra Telesca, Alessandra Vergallito, Anna Vedani, Gaia Locatelli, Benedetta Visiello, Giuseppe Lauria Pinter, Leonor J. Romero Lauro

## Abstract

Chronic Primary Pain (CPP) is a new diagnostic category including chronic pain conditions lacking clinical signs or a clear etiopathogenetic origin. These disorders may share a common neural mechanism known as central sensitization, where nociceptive neurons become hyper-responsive to standard or subthreshold pain stimuli, resulting in pain hypersensitivity. In this context, non-invasive brain stimulation (NIBS) seems a promising tool to improve CPP symptoms by targeting maladaptive brain activity and connectivity. To date, NIBS effects on CPP symptoms remain unexplored. To fill this gap, we conducted a meta-analysis, investigating the effect of NIBS in improving the three core symptoms of CPP, namely pain intensity, emotional distress, and functional disability. Following PRISMA guidelines, we screened four databases up to the end of January 2023. Thirty-five English-written randomized clinical trials were included, comprising 874 participants assigned to the real stimulation condition and 713 to the sham.

Findings highlighted the effect of the real over the sham stimulation in improving CPP core symptoms immediately after the treatment. For pain intensity and functional disability, the improvement persisted also at the one-month follow-up. Meta-regression analyses highlighted that a longer CPP duration reduced the effects of NIBS, while an increased number of sessions was associated with greater pain relief at follow-up.

Taken together, our results suggest that NIBS can effectively alleviate CPP symptoms in the short and medium term. Further research is needed to define standardized NIBS protocols for CPP management and explore whether combining NIBS with other therapeutic interventions can enhance effects duration and efficacy.

## Introduction

Chronic pain is a complex bio-psycho-social condition characterized by the sustained experience of physical pain lasting longer than three months [143]. The disease affects 40% of the global population [31] and seriously interferes with several aspects of patient’s lives, including work or educational careers, physical and physiological functioning, and psychological well-being [122; 123; 130]. The direct and indirect costs associated with chronic pain are estimated at approximately $600 billion annually [140], making the understanding and treatment of the disease a global public health research priority. Despite chronic pain’s widespread impact, therapeutic solutions for the disorder remain largely unsatisfactory [88; 111]. This is especially true for diseases characterized by idiopathic pain, where the cause of the pain is unknown, or for all those diseases where there are no clinical signs that can justify the presence of pain symptoms, for instance, fibromyalgia, persistent idiopathic facial pain, chronic migraine. These conditions have been recently classified by the International Classification of Diseases, 11^th^ Revision (ICD-11) [170] as *Chronic Primary Pain* (CPP), which is opposed to *Chronic Secondary Pain,* where pain is at least initially seen as a symptom secondary to an underlying disease [114]. According to the ICD-11, CPP includes three core symptoms: (i) persistent pain lasting longer than three months and affecting one or more anatomical regions, (ii) significant emotional distress, including anxiety, depressive mood, and the experience of negative emotions like anger and frustration, and/or (iii) functional disability, including the interference in daily activities or social functioning. Crucially, CPP can be diagnosed only when the symptoms cannot be sufficiently accounted for by any other condition.

The new categorization aims to shift the focus from disorder-specific symptoms to the common underlying mechanisms driving these conditions. Although the etiopathogenetic origins of pain in CPP are not fully understood, a widely accepted theory suggests that it may involve central sensitization. Central sensitization is a mechanism in which nociceptive neurons exhibit increased responsiveness to standard or even subthreshold input, leading to pain hypersensitivity [179]. From this perspective, the central nervous system may alter, amplify, or modulate the perception of noxious stimuli. Consequently, pain perception no longer corresponds to the structural characteristics of harmful peripheral stimuli but is shaped by the altered neuroplastic processes. This mechanism has been identified across various chronic pain conditions that are now included in the CPP category [10; 116], thus reinforcing the hypothesis of a common neurophysiological substrate across chronic pain-specific disorders. Neuroimaging studies provided converging evidence of widespread structural and functional anomalies in individuals with specific conditions that are now encompassed in CPP compared to healthy participants. These findings highlight the role of the central nervous system in the experience, development, and maintenance of pain in the disorder [14; 36; 65; 77; 81; 87; 104; 171; 183]. Specifically, structural alterations involve the reduction of gray and white matter volume and cortical thickness in regions implicated in nociceptive processing and in the emotional-affective and regulatory components of the pain network, including the insula, amygdala, (hypo)thalamus, (para)hippocampus, anterior cingulate cortex, and precentral and inferior frontal gyri [24; 65; 78; 104]. Anomalies in functional connectivity within a large-scale brain network have also been reported. These alterations involve the default mode, salience, and central executive networks and extend to regions implicated in cognitive and affective processing (see for reviews Peyron 2019, De Ridder 2022[36; 137]). Such complex and still not fully understood neural mechanisms highlight the multifacet nature of CPP, which includes not only an altered pain experience but also impairments in the emotional and cognitive domains (see Tracey 2009, Kohoutová 2022, Tu 2019, Fallon 2016, Johansson 2024, Telesca et al., 2024[46; 81; 90; 164; 169; 171]).

When considering chronic pain treatment strategy, pharmacological interventions typically represent the first-line clinical choice [124]. Analgesics used across CPP-included disorders are centrally acting drugs, such as nonopioid analgesics, antidepressants with analgesic functions, gabapentinoids, or opioids [3; 15; 42; 86; 93; 128; 144]. However, pharmacological trials yielded unsatisfactory outcomes, with often limited pain relief and a high prevalence of side effects [9; 117]. Therefore, an increasing demand for alternative or add-on multimodal strategies has emerged [23].

Over the past decades, non-invasive brain stimulation (NIBS) techniques gained considerable attention for modulating maladaptive brain activity and connectivity in various psychiatric and neurological conditions [51; 73; 95; 145; 175]. Transcranial magnetic stimulation (TMS) and transcranial electrical stimulation (tES) represent the most frequently used techniques. TMS delivers a strong, short magnetic pulse to the participant’s head, which induces neuronal firing by depolarizing the neuronal membrane suprathreshold [89]. TMS is typically applied through repetitive pulses (rTMS) or patterned protocols (e.g., intermittent or continuous theta burst stimulation, iTBS/cTBS) when applied to induce long-lasting effects. TES includes neuromodulatory techniques in which weak constant or alternating currents are applied to the human brain through scalp electrodes. The most used tES techniques are transcranial direct current stimulation (tDCS) and transcranial alternating current stimulation (tACS). TDCS acts by delivering a constant current (typically 1–2 mA) through two electrodes, a positive (anode) and a negative (cathode) one [120]. The current is too weak to generate action potentials per se; rather, it induces small changes at the membrane potential level, thus influencing neuronal spiking likelihood and, in turn, cortical excitability[22; 141]. Traditionally, the parameters considered to establish whether rTMS and tDCS protocols will produce inhibitory or excitatory effects are frequency for rTMS and polarity for tDCS. Low-frequency rTMS (≤ 1 Hz and continuous theta-burst stimulation) and cathodal tDCS are considered inhibitory protocols. In contrast, high-frequency rTMS (> 5 Hz and intermittent theta-burst stimulation, iTBS) and anodal tDCS are considered excitatory [52; 119]. In the case of tACS, the current alternates at a certain frequency and produces a voltage that cyclically and gradually changes from positive to negative polarity, thus modulating physiologically relevant brain oscillations [132] that affect cortical neurons [43]. Although a detailed discussion of NIBS features and functioning goes beyond the scope of the current work, we want to stress that the outcomes of NIBS in terms of excitability or behavioral modulation are difficult to determine in advance but are the results of a complex interplay among stimulation protocol parameters (intensity, duration, number of sessions), the brain regions stimulated, and their connections within the network, the state of the brain during the stimulation, and individual differences including anatomical, hormonal, and genetic features [115; 133; 174].

The rationale for using NIBS in CPP relies on NIBS’s potential to modulate cortical plasticity through synaptic strengthening (long-term potentiation) or weakening (long-term depression) processes [84; 146] that outlast the stimulation time, thereby rebalancing the previously mentioned maladaptive activity and connectivity patterns [29; 92; 118; 182] involved in pain processing and experience [180]. Recent reviews [21] and meta-analyses [48; 70] on randomized clinical trials discussed NIBS’ application and quantitative effects in specific chronic pain disorders included in CPP. For instance, Hou et al. (2016) investigated the effects of sixteen rTMS and tDCS studies in which NIBS was applied as an add-on treatment to medications in patients with fibromyalgia. Overall, the results suggest that NIBS effectively reduced pain symptoms. Additionally, rTMS over the M1 alleviated fatigue, whereas rTMS over the DLPFC modulated depressive symptoms [70]. Feng et al. (2019) included nine studies investigating the effects of NIBS on patients with migraine. Their results highlight that excitatory stimulation of M1 reduced pain intensity and headache attack frequency after the treatment [48]. Finally, Brighina (2019) discussed the use and impact of tES in patients with fibromyalgia. They reviewed twelve studies and suggested that the delivery of anodal tDCS over the left or bilateral M1 could reduce pain intensity but did not modulate performance in cognitive tasks involving executive functions and attention or symptoms related to the affective domain, including anxiety and depressive symptoms, fatigue, and sleep disturbances. Conversely, the five studies applying tDCS over the left or bilateral DLPFC did not trace changes in pain perception. Still, they highlighted a slight improvement in the previously cited cognitive and affective domains [21].

Taken together, previous data synthesis works suggested the effectiveness of NIBS in treating specific chronic pain conditions, such as fibromyalgia and migraine. However, they overlooked less common pain disorders now classified under the CPP category. According to the central sensitization hypothesis, these disorders may share common neural substrates and mechanisms, and could benefit from NIBS. Furthermore, previous quantitative analyses investigated only pain intensity or headahce attack frequency, neglecting emotional distress and/or the functional impact of the disease on patients, symptoms that are now included in the criteria for CPP diagnosis. Finally, previous works focused on pre to post-treatment outcomes, not including follow-up measurements. Exploring NIBS effects at follow-up would provide crucial information on the durability of treatment [22; 32]. Considering these points, the current work aims to qualitatively characterize and quantitively measure the effectiveness of NIBS as a therapeutic strategy to improve the core symptoms of CPP, including pain intensity, emotional distress, and functional disability. Effects on quality of life were also assessed in a subset of studies including this measure. We considered the outcome measures recorded immediately after the treatment and, when possible, at follow-up. We further explored whether NIBS effects varied as a function of specific stimulation parameters. A final correlation analysis between the obtained effect sizes is included and discussed.

## Methods

The systematic review and meta-analysis followed the Preferred Reporting Items for Systematic Reviews and Meta-Analyses (PRISMA) guidelines [127]. Study quality assessment followed the Cochrane Collaboration’s Risk-of-Bias tool for RCT [66]. Method’s details are reported in Supplementary materials – section A.

### Literature search

Four databases, namely PubMed, Web of Science, Embase, and Scopus, were screened to identify peer-reviewed English-language original studies published up to the end of January 2023 that investigated NIBS in patients suffering from CPP. Keywords related to NIBS were combined with relevant primary pain labels identified using the MeSH Tree Structures in PubMed (Supplementary Materials – Section A). Papers were excluded when: (i) they were written in languages other than English; (ii) they were not original studies (e.g., case reports, reviews, meta-analyses, etc.); (iii) they included non-human samples or (iv) non-idiopathic pain pathologies; (v) applied NIBS without a therapeutic purpose. Moreover, we excluded studies (vi) different from randomized clinical trials (RCT), (viii) not including a sham/placebo condition, (xi) not including the necessary data to run the meta-analysis (see the paragraph “Quantitative analyses” for details). Figure 1 summarizes the screening procedure.

**Figure 1:**
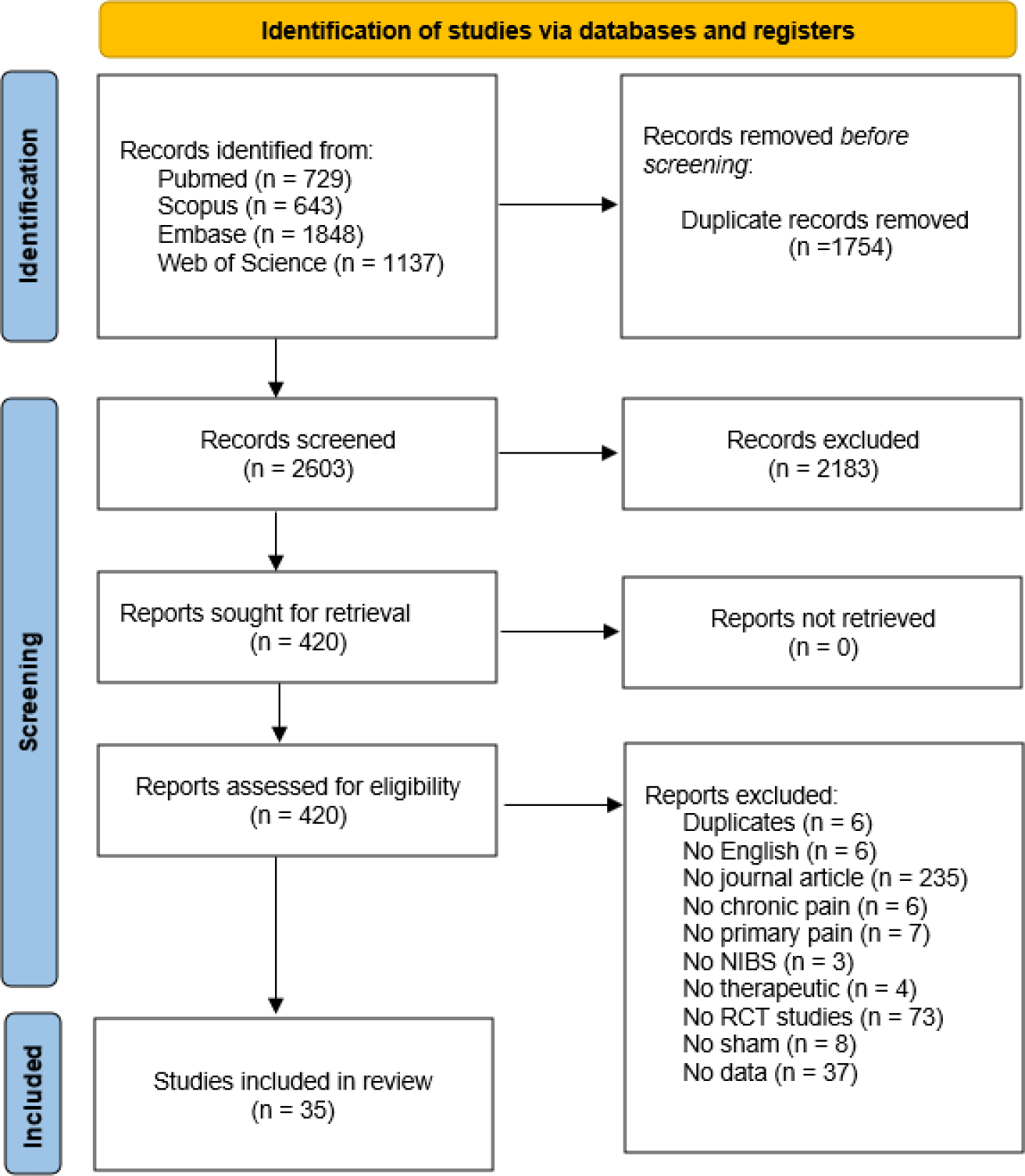
Flowchart of study selection.

### Records screening and data extraction

We used the web and mobile systematic reviews manager Rayyan (https://rayyan.qcri.org/) [126] to run a blind screening process. After removing duplicates, two researchers (A.T. and A.Vedani) independently screened the articles based on titles and abstracts, categorizing the items as “include”, “exclude” or “maybe”, based on the previously outlined criteria. The included articles were screened at the full text using the same blind procedure. When full texts were not available, we contacted the corresponding authors. Conflicting decisions during the title-abstract and full-text screening phases were resolved through consensus. Possible discrepancies were solved with the agreement of all the authors. Then, A.T. and A.Vedani extracted data from the included articles using a structured form that another author checked for consistency and accuracy (A.Vergallito).

The primary outcome measures reflected the three CPP core symptoms as outlined by the ICD-11, namely pain intensity, emotional distress, and functional disability before and after the treatment. Secondary outcome measures included data at follow-up, and participants’ reported quality of life. A detailed description of the included measures is reported in the Supplementary Materials -Section A.

## Quantitative analyses

We extracted relevant information from each study, including NIBS protocol details, sample sizes, and outcome measures. We calculated post to pre-treatment mean differences for real and sham conditions [33]; therefore, negative values represent improved symptoms. We performed analyses in the statistical programming environment R[161], using the metafor package (Viechtbauer, 2010; Viechtbauer & Cheung, 2010). We computed Hedge’s g [63; 64; 176]. Since some studies included more than one effect size, multi-level random effects models were tested [61], and if appropriate, they were reported to handle independence violations [16; 56]. Heterogeneity was assessed using several measures, including the Q statistic (for sampling error variation), I² statistic (for variation not due to sampling error) [67], and prediction intervals (PIs) (range where a future observation is likely to fall) [74]. We used random-effects models to account for heterogeneity due to sampling error and inter-study variance [49]. Subgroup and meta-regression analyses were performed when sufficient data were available [19]. Publication bias was not analyzed when heterogeneity was high (I² ≈ 75%) [61; 142; 173]. A detailed description of the statistical procedure is provided in the Supplementary Materials, section A. Correlations among the effect sizes were explored [178].

## Results

### Studies selection

The research intercepted 4357 records. We removed 2668 as duplicates and 1741 according to the exclusion criteria. We screened 420 full-text articles and after double-blind checking 35 studies were included in the meta-analysis (385 excluded for the following reasons: 6 duplicates, 6 no English, 235 no journal articles, 6 no chronic pain, 7 no primary pain, 3 no NIBS, 4 no therapeutic, 73 no RCT studies, 8 no sham, 37 no available data) (**Figure 1**). **Table 1** includes the key features of the included studies.

**Table 1:** Summary of participants’ characteristics from the included studies.

| TMS protocols |  |  |  |  |  |  |  |  |  |  |  |
| --- | --- | --- | --- | --- | --- | --- | --- | --- | --- | --- | --- |
| Real stimulation group |  |  |  |  |  | Sham stimulation group |  |  |  |  |  |
| Authors | No. of patients | Age | Gender | Education | Illness duration (years) | No. of patients | Age | Gender | Education | Illness duration (years) | Diagnosis (Criteria) |
| Ali AbdElkader et al., 2021 | 16 | n.r. | n.r. | n.r. | 1.83±3 | 11 | n.r. | n.r. | n.r. | 2.83±0.71 | Chronic tension-type headache and chronic migraine (IHS criteria-2004) |
| Altas et al., 2019 | M1: 10, DLPFC: 10 | M1: 46.3±9.01 DLPFC: 47.9±7.89 | F | n.r. | M1: 3±,83; DLPFC: 4.2±1,14 | 10 | 48.2±9.38 | F | n.r. | 3.6±1.43 | Fibromyalgia (ACR 2010) |
| Avery et al., 2015 | 7 | 54.86±7.65 | F | n.r. | 11±4.26 | 11 | 52.09±10 | F | n.r. | 15.64±6.93 | Fibromyalgia (ACR 1990) |
| Bilir et al., 2021 | 10 | 46.70±9.06 | F | 7.25 yrs | 5±[2-5.3] | 10 | 43.80±9.37 | F | 7.25 yrs | 5.5±[4-5.5] | Fibromyalgia (diagnostic criteria 2016) |
| Boyer et al. 2014 | 19 | 49.1±10.6 | F | 47.4% SS | 3.7±4.5 | 19 | 47.7±10.4 | F: 18 M: 1 | 68.4% SS | 3.6±3.8 | Fibromyalgia (ACR 2010) |
| Guinot et al., 2021 | 17 | 46.5±10.4 | F | 9 C, 9 HS | 11.2±10.9 | 19 | 42.8±8.8 | F: 15 M: 4 | 12 C, 7 HS | 9.2±9.6 | Fibromyalgia (ACR 2010) |
| Mhalla et al. 2011 | 20 | 51.8±11.6 | F | n.r. | 13±12.9 | 20 | 49.6±10 | F | n.r. | 14.1±11.9 | Fibromyalgia (ACR 1990) |
| Misra et al., 2013 | 49 | 35.62±10.07 | F: 44 M: 5 | n.r. | 11.19±7.41 | 50 | 35.06±10.38 | F: 44 M: 6 | n.r. | 9.75±7.17 | Chronic migraine (IHS criteria - 1996) |
| Passard et al., 2007 | 15 | 52.6±7.9 | F: 14 M: 1 | n.r. | 8.1±7.9 | 15 | 55.3±8.9 | F | n.r. | 10.9±8.6 | Fibromyalgia (ACR 1990) |
| Picarelli et al., 2010 | 11 | 43.5±12.1 | F: 7<br>M: 5 | n.r. | 6.9±2.9 | 11 | 40.6±9.9 | F: 7<br>M: 4 | n.r. | 6.6±2.7 | Complex Regional Pain Syndrome (CRPS) Type I (IASP)I |
| Tekin et al., 2014 | 27 | 42.4±7.63 | F: 24<br>M: 3 | 8.40±2.85<br>yrs | 10.81±6.31 | 24 | 46.5±8.36 | F: 23<br>M: 1 | 8.13±2.42<br>yrs | 13.33±6.65 | Fibromyalgia (ACR) |
| Todorov et al., 2020 | <u>M1: 38,</u><br><u>IDLPCF:</u><br><u>37</u> | <u>M1:</u><br>40.2±11.05;<br><u>IDLPCF:</u><br>38.7±11.05 | F: 34<br>M: 14 | n.r. | M1: 19.3;<br>IDLPCF:<br>19.2±<br>M1: 8.75;<br>IDLPCF:<br>8.42 | 28 | 38.7±11.05 | F: 22<br>M: 6 | n.r. | 18.5±9,81 | Chronic migraine (IHS criteria - 2013) |
| Umezaki et al. 2016 | 12 | 63.4±10.8 | F:<br>92.9% | n.r. | 5.1±2.7 | 8 | 64.4±8.4 | F:<br>91.7% | n.r. | 5±4.6 | Burning Mouth Syndrome (BMS) (IHS criteria - 2013) |
| YAĞCI et al., 2014 | 12 | 45.25±9.33 | F | n.r. | 4.42±2.43 | 13 | 43±7.63 | F | n.r. | 4.58±2.54 | Fibromyalgia (ACR 1990) |

tES protocols
| Real stimulation condition |  |  |  |  |  | Sham stimulation condition |  |  |  |  | Diagnosis (Criteria) |
| --- | --- | --- | --- | --- | --- | --- | --- | --- | --- | --- | --- |
| Authors | No. of patients | Age | Gender | Education | Illness duration (years) | No. of patients | Age | Gender | Education | Illness duration (years) |  |
| Antal et al., 2011 | 13 | 33.2±10.4 | F: 12<br>M: 1 | n.r. | 15.3±12.1 | 13 | 32.3±12.3 | F: 11<br>M: 2 | n.r. | 12±8.9 | Chronic migraine (IHS criteria - 2004) |
| Arroyo-Fernandez, 2022 | 40 | 50.60±7.01 | F: 38<br>M: 2 |  | 11.85±7.30 | 40 | 49.53±8.74 | F: 39<br>M: 1 | n.r. | 11.30±6.59 | Fibromyalgia (ACR 2010) |
| Caumo, 2022 | 32 | 49.6±9.00 | F | 11.33 ±<br>4.15 | n.r. | 16 | 48.0±6.72 | F | 12.11±4.04 | n.r. | Fibromyalgia (ACR 2016) |
| de Melo et al., 2020 | 11 | n.r. | F | n.r. | n.r. | 11 | n.r. | F | n.r. | n.r. | Fibromyalgia (ACR) |
| Dutra et al., 2020 | 13 | 26.1±3.8 | F | n.r. | n.r. | 11 | 21.0±2.1 | F | n.r. | n.r. | Dysmenorrhea (Primary Dysmenorrhea Consensus Guideline)<sup>3</sup> |
| Fagerlund et al., 2015 | 24 | 49.04±8.63 | F | n.r. | 17.73±7.54 | 24 | 48.17±10.56 | F: 21<br>M: 3 | n.r. | 18.50±11.48 | Fibromyalgia (ACR 1990) |
| Hazime et al., 2017 | <u>PES real +tDCS</u> real: 23<br><u>PES sham + tDCS</u> real: 23 | <u>PES real +tDCS</u> real: 51.9±9.9<br><u>PES sham + tDCS</u> real: 51.3±9.9 | <u>PES real +tDCS</u> real: F: 19, M: 4<br><u>PES sham + tDCS</u> real: F: 18, M: 5 | <u>PES real +tDCS</u> real: Higher Education: 2±8.7<br><u>PES sham + tDCS</u> real: Higher Education: 2±8.7 | 7.6±9 | <u>PES real +tDCS</u> sham: 23<br><u>PES sham +tDCS</u> sham: 23 | <u>PES real +tDCS</u> sham: 51.3 ± 9.9<br><u>PES sham +tDCS</u> sham: 54.1 ± 9.8 | <u>PES real +tDCS</u> sham: F=18 M=5; <u>PES sham +tDCS</u> sham: F: 17, M: 6 | <u>PES real +tDCS</u> sham: Higher education: 2±8.7<br><u>PES sham +tDCS</u> sham: 2±8.7 | 5.8±7.7 | Nonspecific low back pain (medical diagnosis) |
| Hodaj et al., 2022 | 17 | 54.5±10.6 | F: 10 (56%) | n.r. | 27.8±13.5 | 17 | 46.1±14.1 | F: 16 (89%) | n.r. | 24.1±15.7 | Chronic migraine (IHS criteria - 2018) |
| Jiang et al. 2020 | 26 | 39.9±14.2 | F: 12<br>M: 14 | n.r. | 2.1±4.1 | 25 | 44.1±13 | F: 12<br>M: 13 | n.r. | 2.5±3.9 | Low back pain (medical decision) |
| Khedr et al., 2017 | 18 | 31.3±10.99 | F: 17<br>M: 1 | n.r. | 0.51±0.22 | 18 | 33.89±11.18 | F: 17<br>M: 1 | n.r. | 0.50±0.21 | Fibromyalgia (ACR 2010) |
| Lin, 2022 | 18 | 48.3±13.6 | F: 13<br>M: 6 | n.r. | n.r. | 17 | 48.9±12.3 | F: 17<br>M: 2 | n.r. | n.r. | Fibromyalgia (ACR 2016) |
| Luedtke et al., 2015 | 67 | 45±9 | n.r. | n.r. | 8.2±8.8 | 68 | 44±10 | n.r. | n.r. | 7.8±10.4 | Low back pain (European guidelines of non-specific chronic low back pain<sup>2</sup>) |
| Matias, 2022 | 17 | 48.94±13.83 | F | > 1st grade | n.r. | 14 | 49.43±15.14 | F | n.r. | n.r. | Fibromyalgia (ACR 2012) |
| Oliveira et al. 2015 | 16 | 23.8±7.3 | F: 15<br>M: 1 | HS | 2.5±1.4 | 16 | 25.5±6.3 | F: 14<br>M: 2 | HS | 2.8±1.9 | Temporomandibular Disorder<br>(Research Diagnostic Criteria -RDCn.r.TMD) |
| Paula, 2022 | <u>LDN</u>+:21,<br><u>LDN</u> -:22 | <u>LDN</u>+tDCS:<br>49.74±1.97<br><u>Placebo</u>+<br>tDCS:<br>50.57± 2.23 | F | 10.00 ±<br>0.53 | n.r. | <u>LDN</u>+:22,<br><u>LDN</u> -:21 | <u>LDN</u>+sham:<br>48.09±1.56<br><u>Placebo</u>+sham:<br>48.95±2.08 | F | 11.55±0.99 | n.r. | Fibromyalgia<br>(ACR 2016) |
| Riberto et al. 2011 | 11 | 58.3±12.1 | F | 6.3±5.2 | 0.83±0.98 | 12 | 52.4±11.5 | F | 8.9±4.4 | 0.5±0.9 | Fibromyalgia<br>(ACR 1990) |
| Samartin-Veiga, 2022 | <u>M1</u>: 29,<br><u>DLPFC</u>:<br>26<br><u>OIC</u>: 28 | <u>M1</u>:<br>49.38± 8.83<br><u>DLPFC</u>:<br>50.55±8.89<br><u>OIC</u>:<br>50.21±8.20 | F | n.r. | n.r. | 25 | 50.67±8.88 | F | n.r. | n.r. | Fibromyalgia<br>(ACR 2010) |
| Segal, 2021 | 9 | n.r. | n.a | n.r. | n.r. | 10 | n.r. | n.r. | n.r. | n.r. | Phantom Limb<br>(Mean daily phantom pain intensity ≥ 4) |
| Serrano, 2022 | 24 | 49.18 ± 8.63 | F | 10.23 ±<br>3.75 | n.r. | 12 | 46.09±11.34 | F | 12.64±5.07 | n.r. | Fibromyalgia<br>(ACR 2016) |
| To et al. 2017 | <u>OC</u>: 15<br><u>FC</u>: 11 | <u>OC</u>:<br>47.13±10.01;<br><u>FC</u>:<br>47.81±10.17 | <u>OC</u>:<br>F:12,<br>M: 3;<br><u>FC</u>: F:<br>10, M:<br>1 | n.r. | n.r. | 16 | n.r. | F:14<br>M: 2 | n.r. | n.r. | Fibromyalgia<br>(ACR 1990) |
| Volz et al., 2016 | 10 | 40.6±12.5 | F: 7<br>M: 3 | n.r. | 10±8.9 | 10 | 34.4±13.2 | F: 6<br>M: 4 | n.r. | 7±4.7 | Inflammatory bowel syndrome<br>(Medical report, discharge letter, and outpatient center records) |

Notes: n.r. = not reported; F= female; M = male; C= College, HS = High School; SS = Secondary School; yrs = years. M1 = motor primary cortex; DLPFC = dorsolateral prefrontal cortex; LDN = Low-Dose Naltrexone; tDCS = transcranial Direct Current Stimulation; OIC = operculo-insular cortex; OC = occipital cortex; FC = frontal cortex; IDLPFC = left dorsolateral prefrontal cortex; PES = Peripheral Electrical Stimulation.

### Study quality assessment

The quality assessment results are reported in **Figure 2**. We calculated the percentage of high-risk judgments to obtain a quality score for each study. Only 5 studies (14.2%) were assessed as low risk, 7 (20%) were assessed as high risk, and the other were evaluated as having some concerns (65.8%). The most omitted domain was *deviation from the intended intervention*, as many studies experienced dropouts and did not account for intention-to-treat analysis. The most respected domains were *missing outcome data* assessing the completeness of outcome data, and *measurements of the outcome*, suggesting that researchers were overall able to completely assess outcome data, and that outcomes were unlikely to be influenced by the knowledge of intervention administered. Since only randomized controlled trials were selected, the domain of randomization was respected, although for some there were some concerns, mainly related to the allocation sequence [148; 168], particularly in some studies it was unclear whether the allocation sequence was concealed until participants were enrolled and assigned to interventions [5; 85; 148] or there were baseline differences between intervention groups [13; 41].

**Figure 2:**
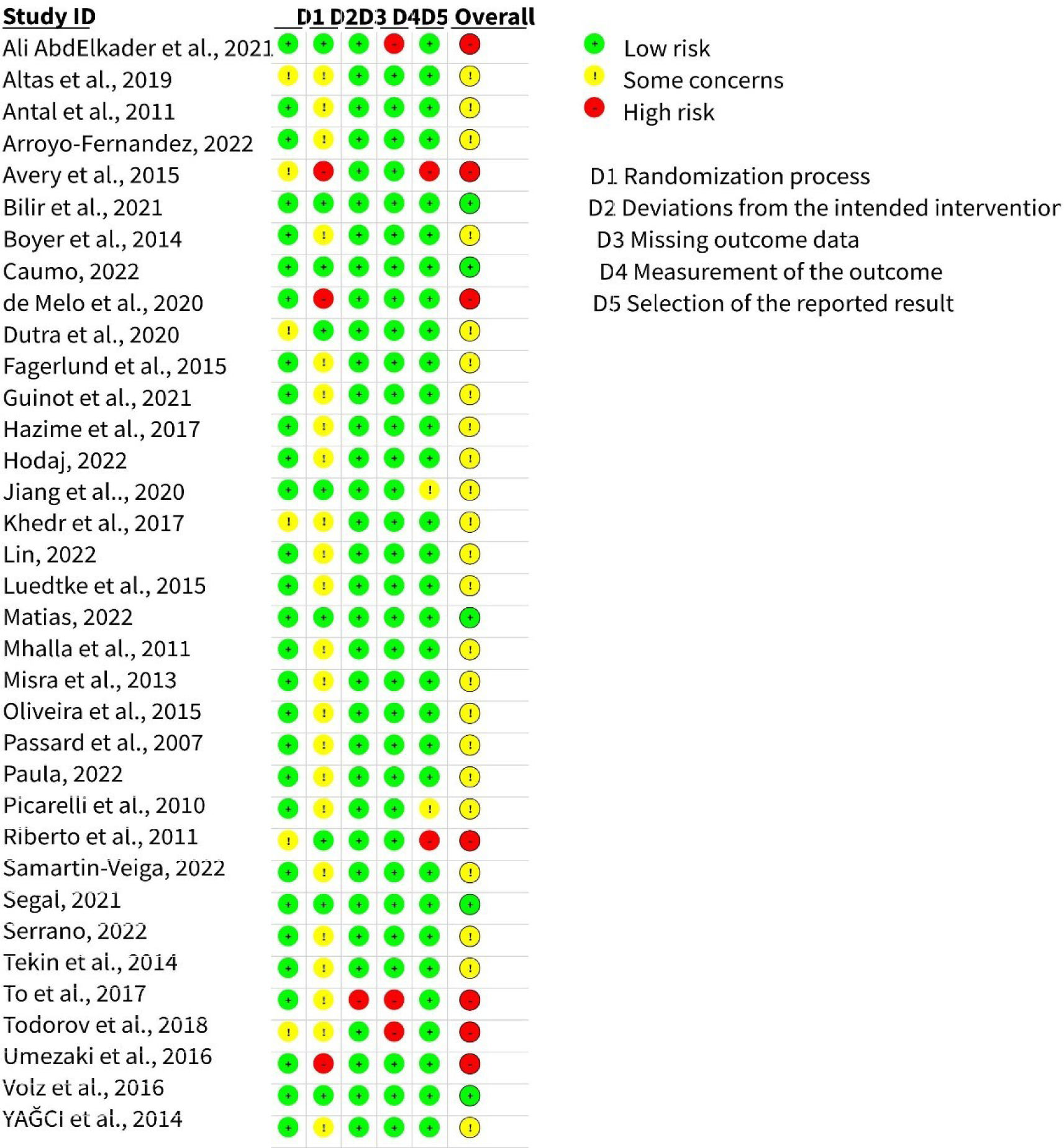
The graphical representation of risk of bias assessed for the 35 studies included in the meta-analysis.

### Patients’ demographical and clinical features

35 studies were included in the meta-analysis, involving 874 patients with CPP assigned to stimulation groups and 713 to control sham groups. **Table 1** shows details on patients’ characteristics and diagnoses. Participants ranged from 21 to 64 years, with a mean age of 45 ± 9.11 years for the group receiving the real stimulation and 44.1 ± 8.62 years for the group receiving the sham. In most studies, the number of females was greater than males, in line with studies reporting the higher prevalence of women suffering from CPP [69; 164]. Moreover, 16 studies included exclusively women [5; 13; 18; 20; 25; 34; 35; 41; 45; 60; 105; 107; 148; 151; 157]. Sex differences in chronic pain are widely debated [50; 59].

The studies’ inclusion criteria required participants to be adults (over 18 years old) with a medical diagnosis of chronic pain, defined as pain lasting more than 3 months [143]. Exclusion criteria generally concerned contraindications to NIBS, neurological and psychiatric comorbidities, pregnancy and breastfeeding in women, a history of substance abuse, and a history of surgery or implanted devices. The studies concerned several chronic pain conditions, now included in the CPP category. Specifically, 21 studies included patients with Chronic Widespread Pain (i.e., fibromyalgia), 7 Chronic Primary Headache or Orofacial Pain (chronic migraine [2; 7; 68; 108; 168], temporomandibular disorder [125], burning mouth syndrome [172]), 4 Chronic primary musculoskeletal pain (chronic low back pain [62; 79; 98], phantom limb pain [155], 2 Chronic Primary visceral pain (irritable bowel syndrome [177], primary dysmenorrhea [41]), and 1 Complex regional pain syndrome [138].

The summary of participants’ characteristics (demographics and diagnosis) is showed in **Table 1**, dividing the studies in TMS and tDCS stimulation groups. NIBS protocols’ features are described in two separate tables: **Table 2** will be dedicated to TMS descriptives and **Table 3** to tES descriptives.

**Table 2:** Qualitative data on TMS protocols used in the included studies. All protocols had repetitive TMS stimulation, only one had a single-pulse stimulation protocol.

| Study | Sessions number | Frequency and intensity* | Pulses number | Duration (minutes) | Protocol type | Coil type | Target region | Therapeutic Strategy<sup>1</sup> |
| --- | --- | --- | --- | --- | --- | --- | --- | --- |
| **Ali AbdElkader et al., 2021** | 12 | 5 Hz, 90% | 1000 | 5 | Excitatory | Figure-of-8 coil | LDLPFC | n.r. |
| **Altas et al., 2019** | 15 | 10 Hz, 90% | 1200 | 30 | Excitatory | Figure-of-8 coil | i) LM1,<br>ii) LDLPFC | Add-on |
| **Avery et al., 2015 #** | 15 | 10Hz, 120% | 3000 | 37,5 | Excitatory | Not specified | LDLPFC | Add-on |
| **Bilir et al., 2021** | 14 | 10Hz, 90% | 1500 | 15 | Excitatory | Figure-of-8 coil | LDLPFC | Add-on |
| **Boyer et al. 2014** | 14 | 10Hz, 90% | 2000 | 20 | Excitatory | Figure-of-8 coil | LM1 | Add-on |
| **Guinot et al., 2021** | 16 | 10 Hz, 80% | 2000 | 20 | Excitatory | Not specified | Dominant M1 | Augmentative |
| **Mhalla et al. 2011** | 14 | 10Hz, 80% | 1500 | 15 | Excitatory | Figure-of-8 coil | LM1 | Add-on |
| **Misra et al., 2013** | 3 | 10 Hz, 70% | 600 | 15 | Excitatory | Figure-of-8 coil | LM1 | Monotherapy |
| **Passard et al., 2007** | 10 | 10 Hz, 80% | 2000 | 25 | Excitatory | Figure-of-8 coil | LM1 | Add-on |
| **Picarelli et al., 2010** | 10 | 10 Hz, 100% | 2500 | 30 | Excitatory | Figure-of-8 coil | M1 | Add-on |
| **Tekin et al., 2014** | 10 | 10Hz, 100% | 1500 | 10 | Excitatory | Figure-of-8 coil | LM1 | Monotherapy |
| **Todorov et al., 2020** | 5 | 15 Hz, 70% | 1200 | 8 | Excitatory | Figure-of-8 coil | M1<br>LDLPFC | Add-on |
| **Umezaki et al., 2016** | 10 | 10Hz, 110% | 3000 | 15 | Excitatory | Figure-of-8 coil | LDLPFC | Add-on |
| **YAĞCI et al., 2014** | 10 | 1Hz, 90% | 1200 | 15 | Inhibitory | Parabolic coil | LM1 | Add-on |

<sup>1</sup> : monotherapy = when patients did not receive additional medications or treatments; add-on = when added to existing interventions (i.e. pharmacotherapy); augmentative approach = when other pharmacological, behavioral, or psychological interventions were combined to the stimulation protocol [145].
\* Intensity is reported as percentage of the resting motor threshold. # single-pulse TMS protocol.
**Note: n.r. = not reported. LDLPFC = left dorsolateral prefrontal cortex; LM1 = left primary motor cortex; M1 = primary motor cortex.**

**Table 3:**
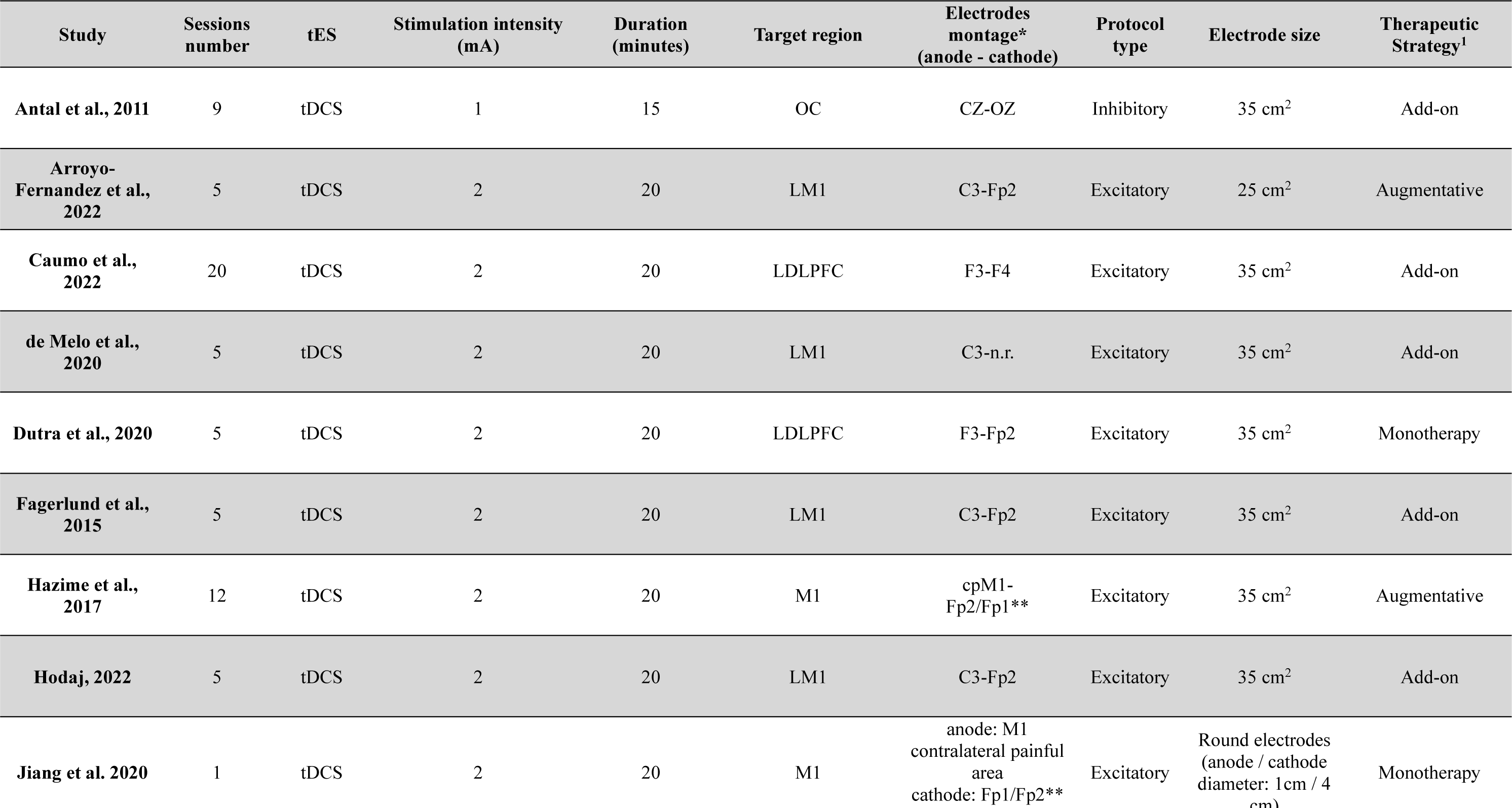
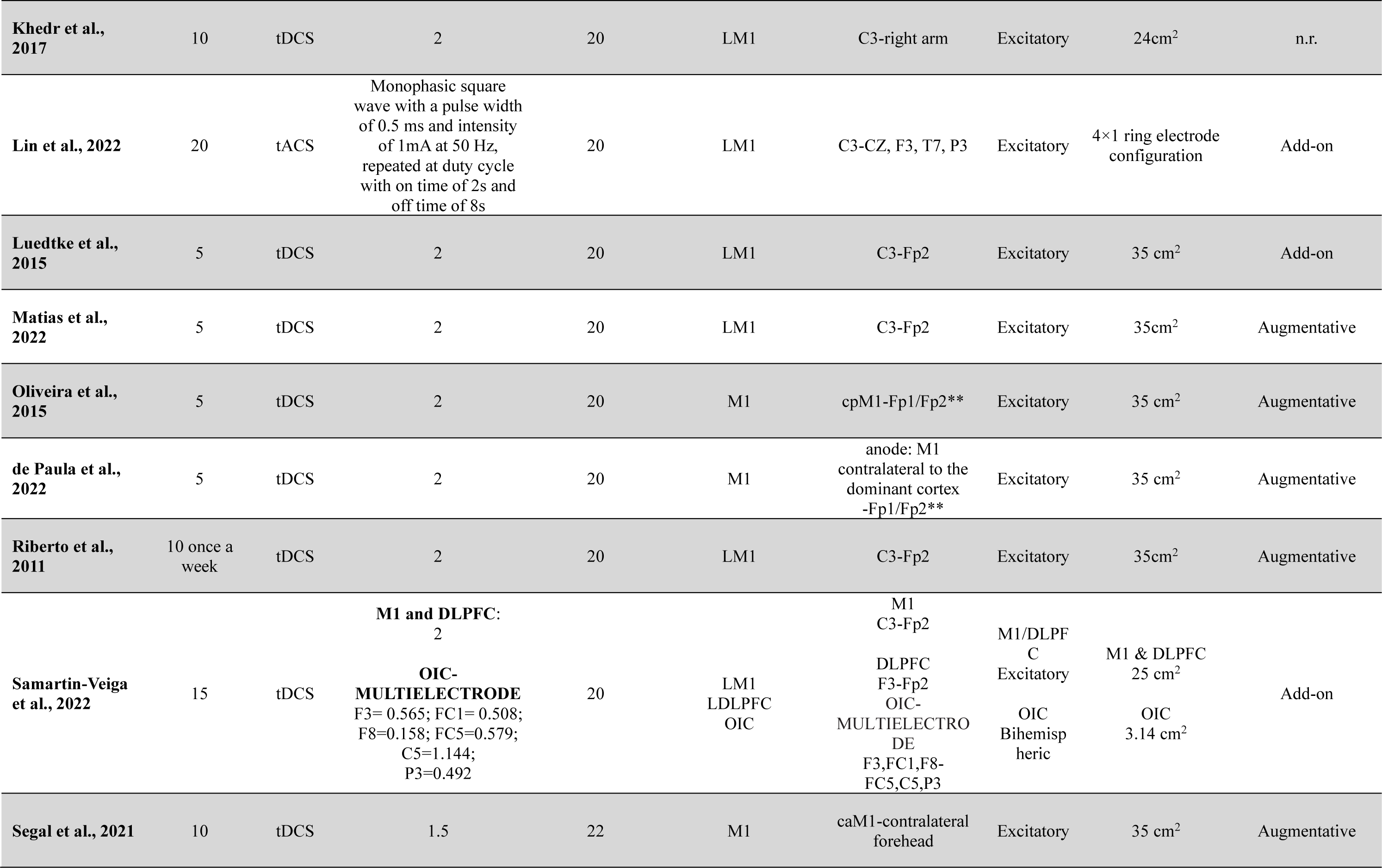
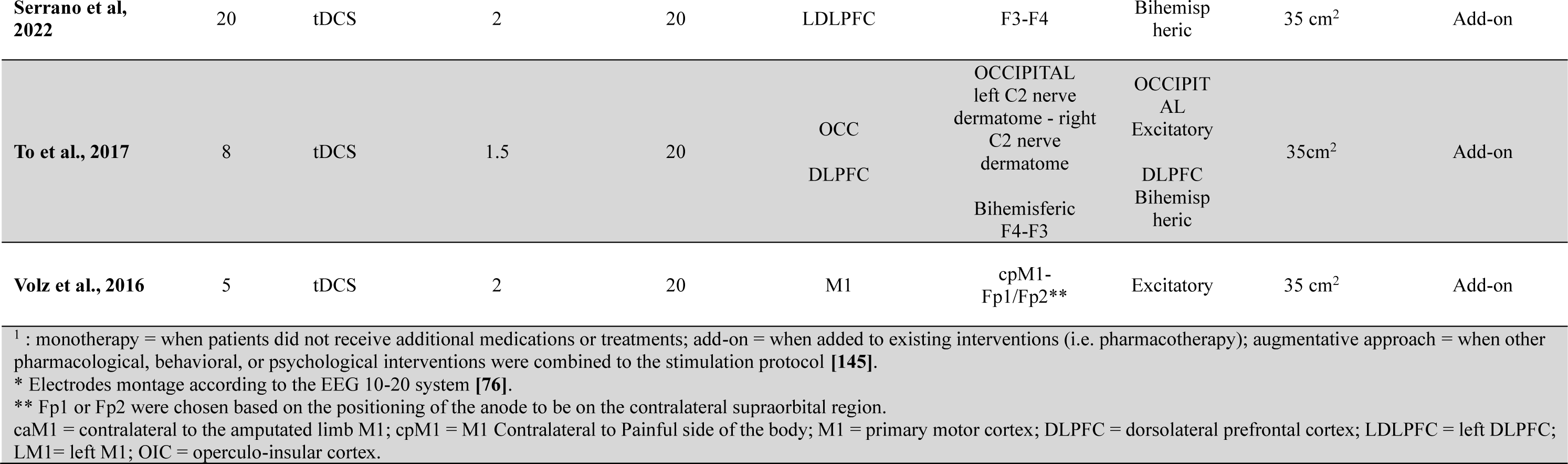
Qualitative data on tES protocols used in the included studies.

| Study | Sessions number | tES | Stimulation intensity (mA) | Duration (minutes) | Target region | Electrodes montage* (anode - cathode) | Protocol type | Electrode size | Therapeutic Strategy<sup>1</sup> |
| --- | --- | --- | --- | --- | --- | --- | --- | --- | --- |
| Antal et al., 2011 | 9 | tDCS | 1 | 15 | OC | CZ-OZ | Inhibitory | 35 cm<sup>2</sup> | Add-on |
| Arroyo-Fernandez et al., 2022 | 5 | tDCS | 2 | 20 | LM1 | C3-Fp2 | Excitatory | 25 cm<sup>2</sup> | Augmentative |
| Caumo et al., 2022 | 20 | tDCS | 2 | 20 | LDLPFC | F3-F4 | Excitatory | 35 cm<sup>2</sup> | Add-on |
| de Melo et al., 2020 | 5 | tDCS | 2 | 20 | LM1 | C3-n.r. | Excitatory | 35 cm<sup>2</sup> | Add-on |
| Dutra et al., 2020 | 5 | tDCS | 2 | 20 | LDLPFC | F3-Fp2 | Excitatory | 35 cm<sup>2</sup> | Monotherapy |
| Fagerlund et al., 2015 | 5 | tDCS | 2 | 20 | LM1 | C3-Fp2 | Excitatory | 35 cm<sup>2</sup> | Add-on |
| Hazime et al., 2017 | 12 | tDCS | 2 | 20 | M1 | cpM1-Fp2/Fp1** | Excitatory | 35 cm<sup>2</sup> | Augmentative |
| Hodaj, 2022 | 5 | tDCS | 2 | 20 | LM1 | C3-Fp2 | Excitatory | 35 cm<sup>2</sup> | Add-on |
| Jiang et al. 2020 | 1 | tDCS | 2 | 20 | M1 | anode: M1<br>contralateral painful area<br>cathode: Fp1/Fp2** | Excitatory | Round electrodes (anode / cathode diameter: 1cm / 4 cm) | Monotherapy |
| **Khedr et al., 2017** | 10 | tDCS | 2 | 20 | LM1 | C3-right arm | Excitatory | 24cm<sup>2</sup> | n.r. |
| **Lin et al., 2022** | 20 | tACS | Monophasic square wave with a pulse width of 0.5 ms and intensity of 1mA at 50 Hz, repeated at duty cycle with on time of 2s and off time of 8s | 20 | LM1 | C3-CZ, F3, T7, P3 | Excitatory | 4×1 ring electrode configuration | Add-on |
| **Luedtke et al., 2015** | 5 | tDCS | 2 | 20 | LM1 | C3-Fp2 | Excitatory | 35 cm<sup>2</sup> | Add-on |
| **Matias et al., 2022** | 5 | tDCS | 2 | 20 | LM1 | C3-Fp2 | Excitatory | 35cm<sup>2</sup> | Augmentative |
| **Oliveira et al., 2015** | 5 | tDCS | 2 | 20 | M1 | cpM1-Fp1/Fp2** | Excitatory | 35 cm<sup>2</sup> | Augmentative |
| **de Paula et al., 2022** | 5 | tDCS | 2 | 20 | M1 | anode: M1 contralateral to the dominant cortex -Fp1/Fp2** | Excitatory | 35 cm<sup>2</sup> | Augmentative |
| **Riberto et al., 2011** | 10 once a week | tDCS | 2 | 20 | LM1 | C3-Fp2 | Excitatory | 35cm<sup>2</sup> | Augmentative |
| **Samartin-Veiga et al., 2022** | 15 | tDCS | M1 and DLPFC: 2<br>OIC-MULTIELECTRODE<br>F3= 0.565; FC1= 0.508;<br>F8=0.158; FC5=0.579;<br>C5=1.144;<br>P3=0.492 | 20 | LM1<br>LDLPFC<br>OIC | M1<br>C3-Fp2<br>DLPFC<br>F3-Fp2<br>OIC-MULTIELECTRODE<br>F3,FC1,F8-FC5,C5,P3 | M1/DLPFC<br>Excitatory<br>OIC<br>Bihemispheric | M1 & DLPFC<br>25 cm<sup>2</sup><br>OIC<br>3.14 cm<sup>2</sup> | Add-on |
| **Segal et al., 2021** | 10 | tDCS | 1.5 | 22 | M1 | caM1-contralateral forehead | Excitatory | 35 cm<sup>2</sup> | Augmentative |
| **Serrano et al, 2022** | 20 | tDCS | 2 | 20 | LDLPFC | F3-F4 | Bihemispheric | 35 cm<sup>2</sup> | Add-on |
| **To et al., 2017** | 8 | tDCS | 1.5 | 20 | OCC<br>DLPFC | OCCIPITAL<br>left C2 nerve<br>dermatome - right<br>C2 nerve<br>dermatome<br>Bihemispheric<br>F4-F3 | OCCIPITAL<br>Excitatory<br>DLPFC<br>Bihemispheric | 35cm<sup>2</sup> | Add-on |
| **Volz et al., 2016** | 5 | tDCS | 2 | 20 | M1 | cpM1-Fp1/Fp2** | Excitatory | 35 cm<sup>2</sup> | Add-on |

<sup>1</sup> : monotherapy = when patients did not receive additional medications or treatments; add-on = when added to existing interventions (i.e. pharmacotherapy); augmentative approach = when other pharmacological, behavioral, or psychological interventions were combined to the stimulation protocol [145].
\* Electrodes montage according to the EEG 10-20 system [76].
\*\* Fp1 or Fp2 were chosen based on the positioning of the anode to be on the contralateral supraorbital region.
caM1 = contralateral to the amputated limb M1; cpM1 = M1 Contralateral to Painful side of the body; M1 = primary motor cortex; DLPFC = dorsolateral prefrontal cortex; LDLPFC = left DLPFC; LM1= left M1; OIC = operculo-insular cortex.

### NIBS protocols

Fourteen of the 35 studies used the TMS, and twenty used a tES protocol (19 tDCS and 1 tACS). **Tables 2** and **3** summarize protocol features and main results of TMS and tES, respectively. A more detailed description is reported in the Supplementary Materials – Section B.

### TMS protocols

RTMS was applied in all studies except Avery et al., 2015 [13], who delivered single-pulse stimulation. 13 studies chose an excitatory strategy, targeting the left dorsolateral prefrontal cortex (DLPFC, 4 studies [2; 13; 18; 172]) or the left motor cortex (M1, 6 studies [20; 107; 108; 129; 162; 181]. Two studies targeted the left M1 and the left DLPFC [5; 168] in two independent groups. Finally, one study targeted the M1 cortex corresponding to the dominant hand [60].

Stimulation frequencies ranged from 5 to 15 Hz, with duration ranging from 5 to 40 minutes and a total number of pulses per session between 600 and 3000. The stimulation intensity ranged between 70% [108; 168] and 120% [13] of the resting motor threshold (rMT), which is the minimum intensity required to evoke a motor response in the target muscle greater than 50 μV in at least 50% of trials [150]. The number of sessions ranged from 3 to 20. Only one study applied an inhibitory protocol [181], delivering 1Hz stimulation to the left M1, with an intensity of 90% of the rMT.

No major adverse reactions were reported. Mild and transient side effects like headache, tinnitus, dizziness, and neck pain were reported in six studies (∼ 43%) with no significant difference between the real stimulation group and the sham stimulation group. Sham stimulation typically consisted of using a sham/placebo coil to mimic the stimulation noise, different inclinations of the coil, or a reduced stimulator output intensity.

### TES protocols

Of the 20 studies applying tDCS, 17 applied anodal stimulation^1^. 9 studies targeted the left M1 [11; 34; 45; 68; 98; 105; 148; 151], 5 the M1 contralateral to the painful area [62; 79; 125; 155; 177], and 3 the M1 corresponding to the hand-dominant hemisphere [35; 62; 79]. The two studies using an extracephalic montage applied the anode over M1 and the cathode on the contralateral arm [85; 167]. Two studies targeted the DLPFC bilaterally, delivering anodal stimulation over the left and cathodal over the right (F3 with F4 cathode per EEG 10-20 IS) [25; 157]. Interestingly, both studies used a home-based protocol. A study included three arms receiving real stimulation, including left M1, left DLPFC, and operculo-insular cortex (OIC), with 15 sessions lasting 20 minutes, and 2mA of stimulation intensity [151]. One study applied cathodal stimulation over the occipital cortex [7].

TDCS intensity varied from 1 to 2 mA with electrode sizes ranging from 25 cm^2^ to 35 cm^2^. The number of sessions varied from 1 to 20, with a session duration varying between 15 and 22. Only one study used tACS, targeting the left M1 with monophasic square waves.

In line with previous safety guidelines [6], no major adverse reactions were reported. Fourteen (∼66%) studies suggested mild and transient side effects, including headache, neck pain, tingling, and skin redness. Sham stimulations were typically delivered using the same devices and identical electrode placement. In sham, the current was turned off after a few seconds of stimulation, typically 30 seconds [54].

## Quantitative results

### Primary outcome measure: pain intensity change score before and after the intervention

Thirty-three effect sizes were computed. The best-fitting model was the reduced one (see **Table S5** - Supplementary materials for details). The meta-analysis results are summarized in the forest plot (**Figure 3**). The random effects model showed a significant effect *g* = - 0.53, 95% CI [-0.76, - 0.31], z = - 4.58, p <.001, suggesting that real stimulation has a moderate impact on reducing the pain experienced by participants after the treatment. The meta-analysis also revealed substantial heterogeneity between studies Q (32) = 122.57, p < .001, τ^2^ = 0.32 (SE = 0.12), and I^2^ = 73.89% [65.06; 88.66]. PIs [-1.70, 0.64] crossed the zero, thus indicating that null or detrimental intervention effects cannot be ruled out for future studies. The Baujat plot inspection **(Figure 4)** suggested that the effect size 16 (Dutra et al., 2020) greatly contributed to the heterogeneity of the statistical analysis. The influence analysis confirmed this study as an influential case. The effect size removal, however, did not change the overall significance: g = - 0.48, 95% CI [-0.69, -0.26], z = -4.37, p <.001.

**Figure 3.**
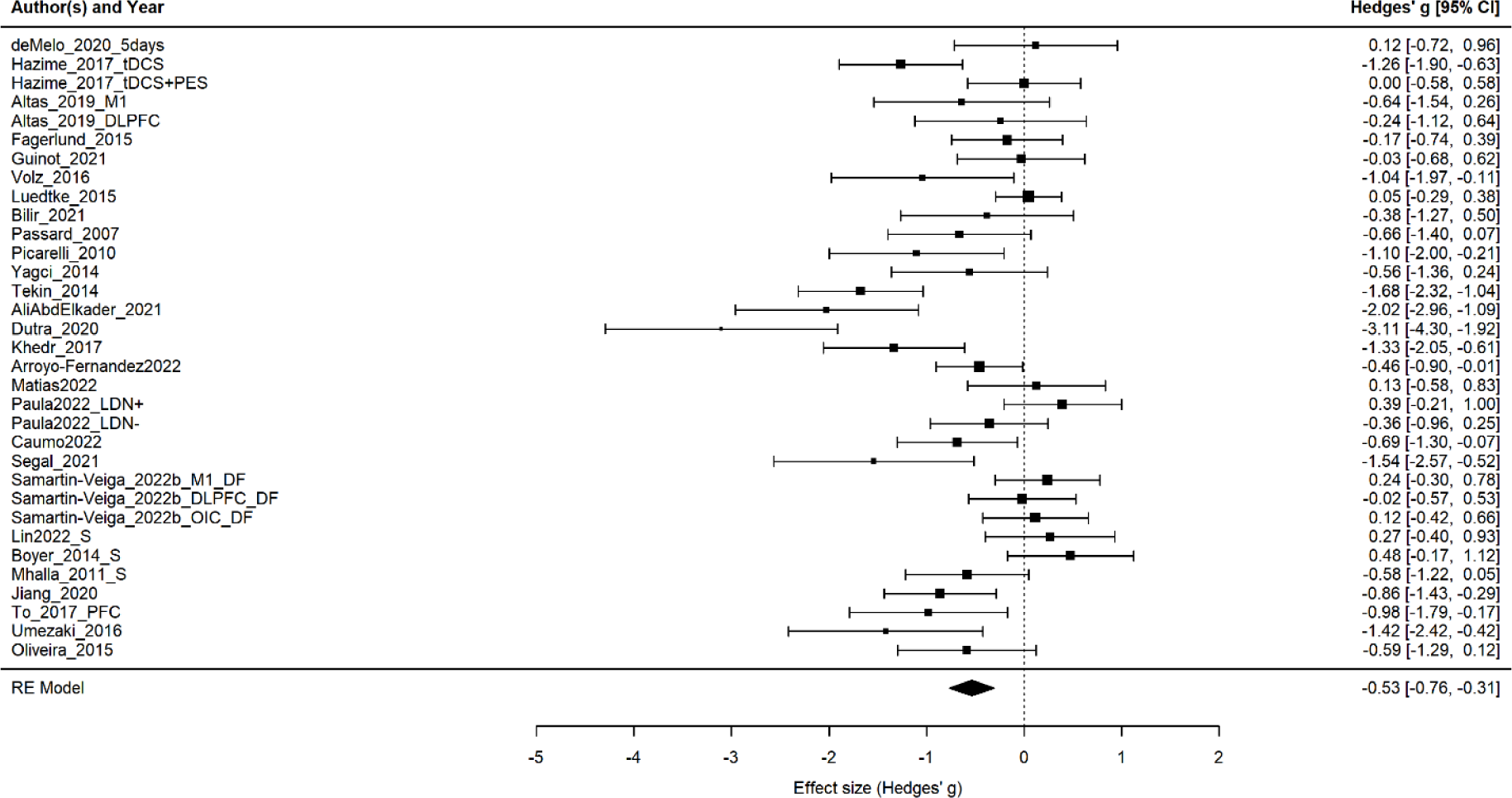
Forest plot of the effect size of noninvasive brain stimulation on pain intensity change score. CI = confidence interval.

**Figure 4.**
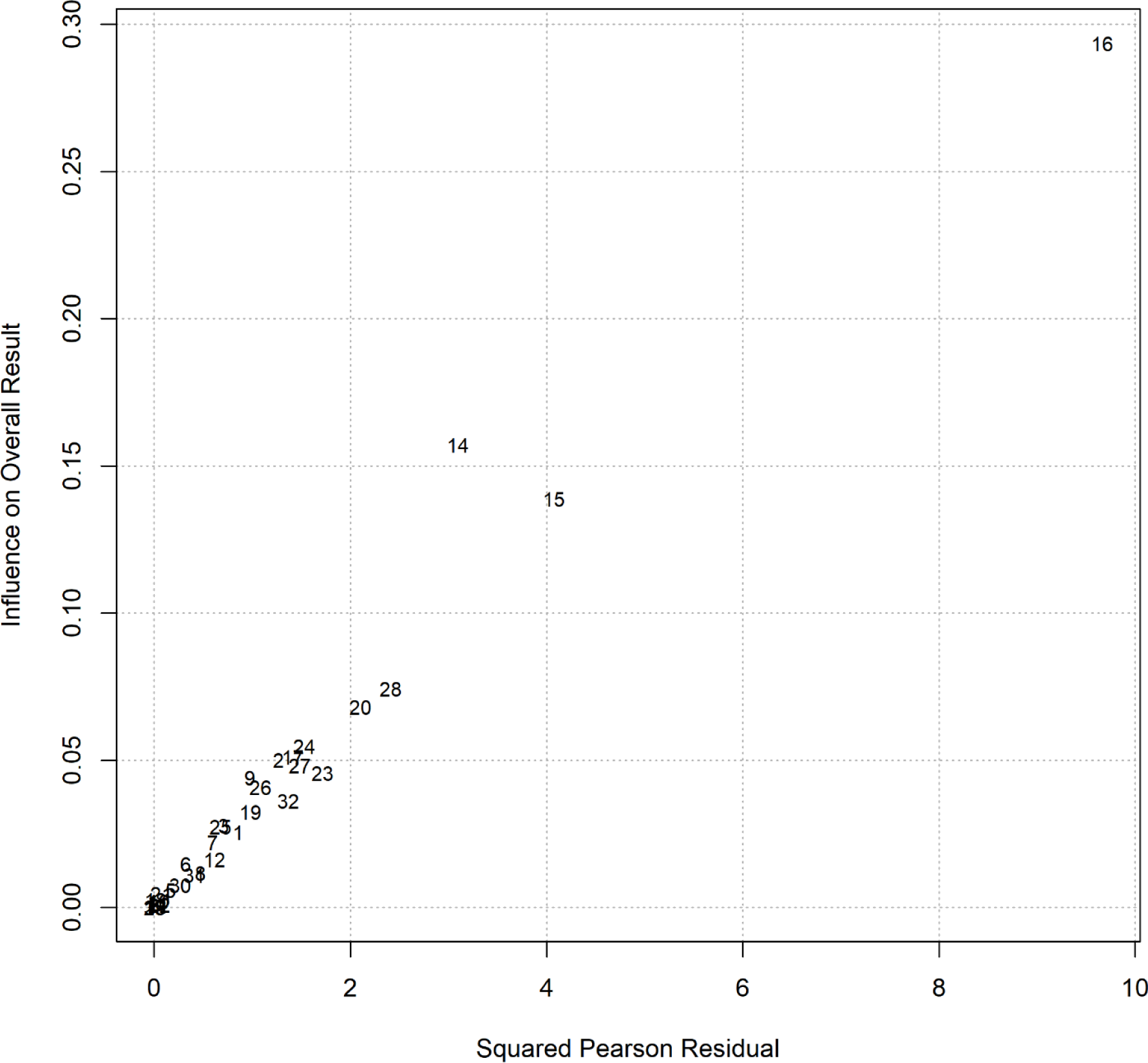
Baujat plot of studies distribution considering pain intensity as the outcome measure.

The included moderators and covariates did not highlight significant patterns (see **Tables S6** and **S7** for details). Publication bias was not explored due to the high heterogeneity.

### Primary outcome measures: emotional distress change score before and after the intervention

The multi-level model was the best-fitting one (see **Table S8**), including 17 studies comprising 32 effect sizes, mostly quantifying depressive and anxiety symptoms. By analyzing the studies’ heterogeneity, however, it was clear that all the variability was explained at the cluster level, corresponding to the between-studies heterogeneity variance computed in conventional two-level meta-analyses. Since the three-level model did not represent the variability in our data, we selected the reduced one (see Supplementary Materials section B for a detailed discussion) [61].

The meta-analysis results are summarized in the forest plot (**Figure 5**). The random effects model was significant *g* = - 0.18, 95% CI [-0.32, - 0.04], z = - 2.48, p = .013. This result suggests that real stimulation may have a small but significant impact on reducing the emotional distress experienced by participants following the real stimulation. The meta-analysis also revealed moderate heterogeneity between studies Q (31) = 53.52, p = .007, τ^2^ = 0.07 (SE = 0.04), and I^2^ = 42.08% [12.57; 75.10]. PIs [-0.73, 0.37] crossed the zero, thus indicating that null or ‘negative’ treatment effects cannot be ruled out for future studies. The Baujat plot inspection (**Figure 6**) suggested that the effect size 13 (Khedr et al., 2017 – anxiety) greatly contributed to the heterogeneity of the statistical analysis, and the influence analysis confirmed it. The effect size removal, however, did not change the overall significance: g = - 0.13, 95% CI [-0.26, -0.01], z = -2.11, p =.035, but it reduced the heterogeneity Q (30) = 39.43, p = .116, τ^2^ = 0.03 (SE = 0.03) and I^2^ = 23.92% [0; 66.72] (low heterogeneity), and PIs [-0.51, 0.24]. Considering the publication bias, the modified Egger test showed no asymmetry b = 0.73, 95% CI [- 0.83, 2.29], z = - 1.17, p = .243.

**Figure 5.**
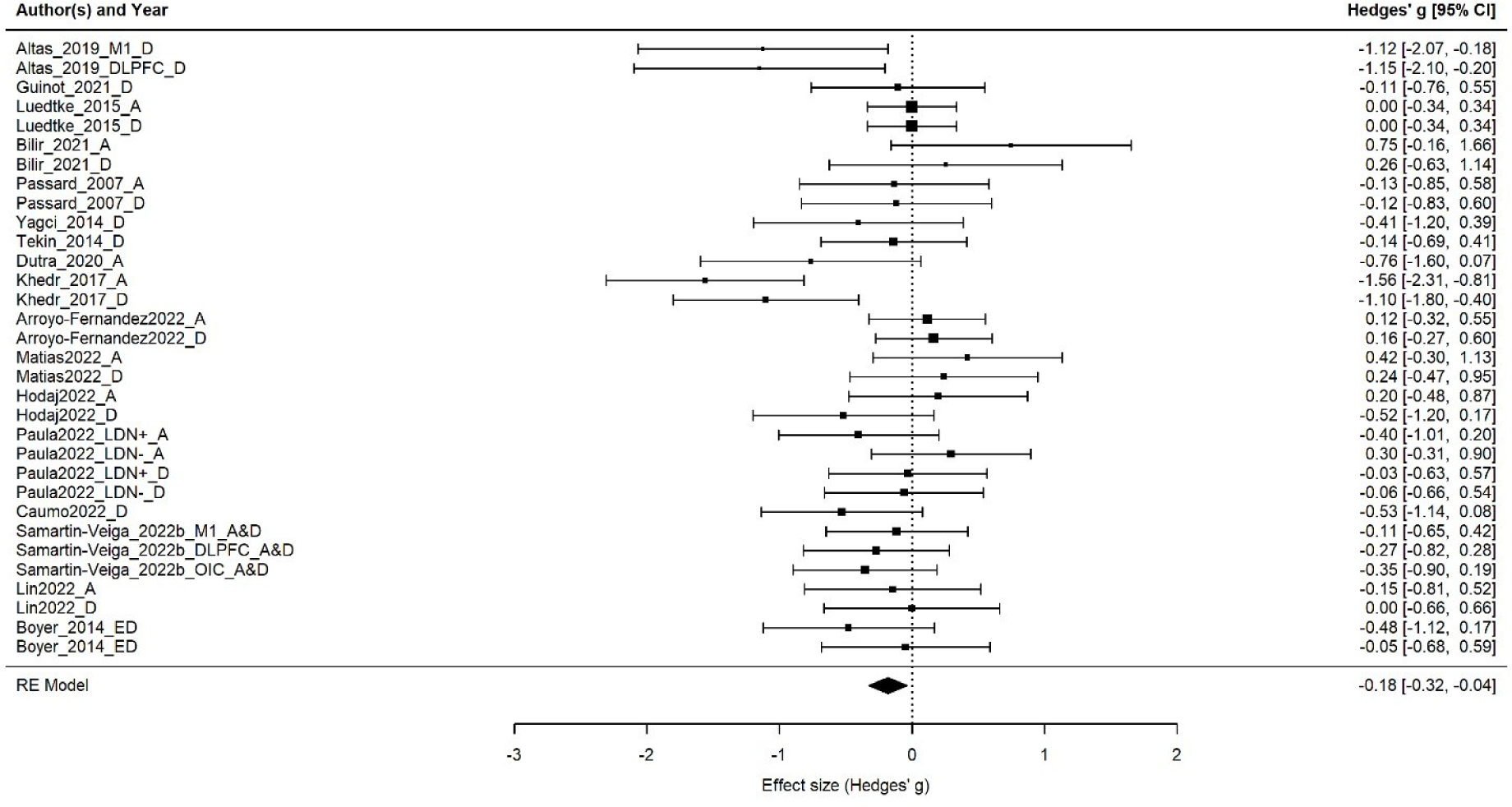
Forest plot of the effect size of noninvasive brain stimulation on emotional distress change score. CI = confidence interval.

**Figure 6.**
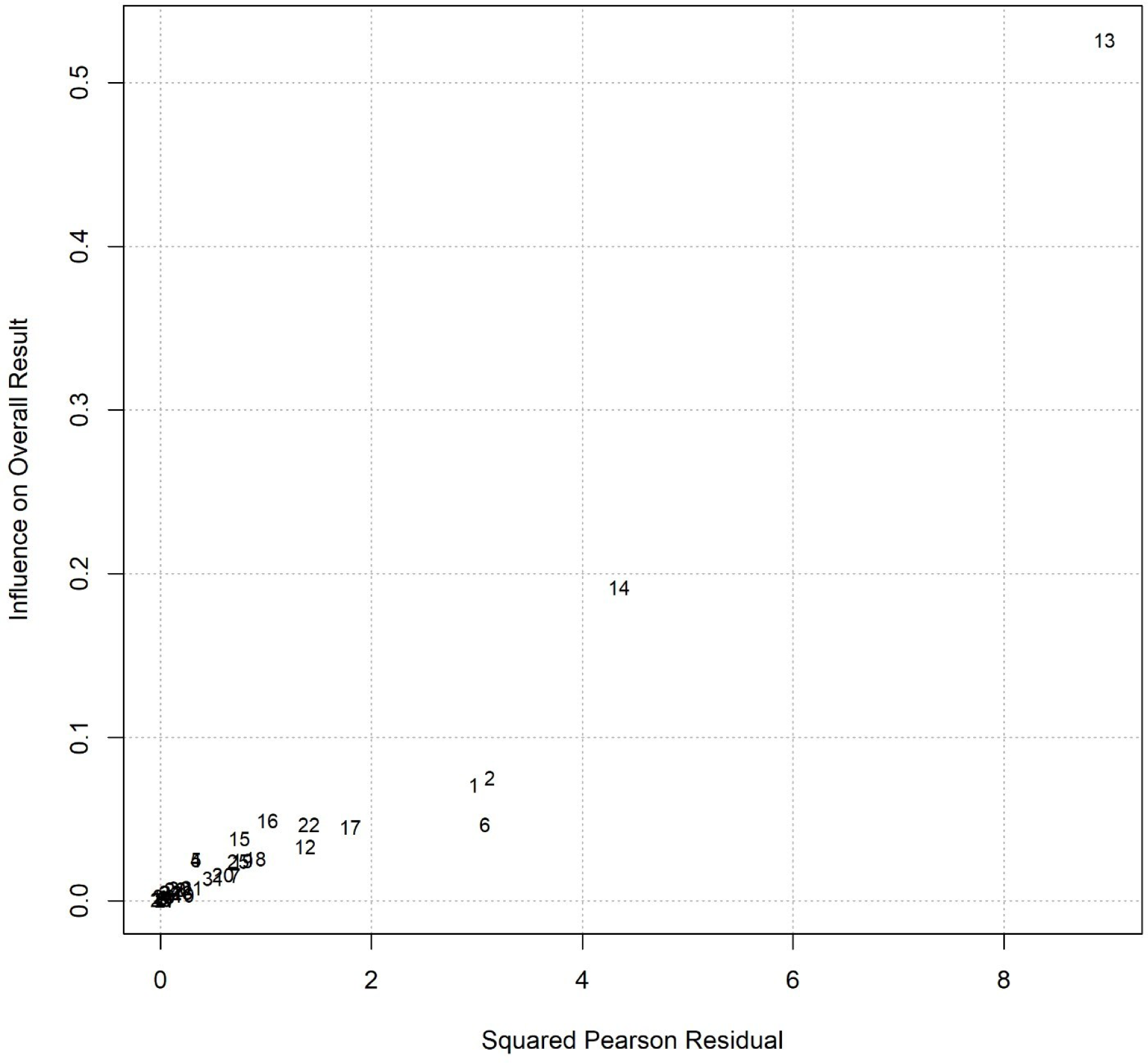
The funnel plot graphically represents the publication bias of the included studies.

Subgroup analysis did not highlight differences (see **Table S8**). The meta-regression analysis indicated a trend toward significance (p=.065) for the illness duration predictor (see **Table S9**). It indicates that for each additional year of illness, the effectiveness of real stimulation in reducing emotional distress decreases, making it less effective as the duration of CPP increases.

### Primary outcome measures: functional disability

Twenty-two effect sizes were computed. The best-fitting model was the reduced one. The meta-analysis results are summarized in the forest plot (**Figure 7**). The random effects model was significant *g* = - 0.31, 95% CI [-0.44, - 0.18], z = - 4.76, p <.001. This result suggests that real stimulation has a small but significant impact on reducing the functional disability rated by participants after the stimulation. The meta-analysis also revealed low heterogeneity between studies Q (21) = 19.99, p = .522, τ^2^ = 0 (SE = 0.03), and I^2^ = 0% [0; 53.73]. PIs [-0.45, -0.18] lay completely on the “negative” side. Therefore, we could expect the intervention to be beneficial across the contexts we studied. Baujat plot inspection (**Figure 8**) suggested the effect sizes 6 and 8 [98; 129] as potential outliers. The influence analysis confirmed only Luedke et al. (2015) as an influential case. The effect size removal, however, did not change the overall significance: g = - 0.34, 95% CI [-0.48, -0.20], z = -4.76, p <.001.

**Figure 7.**
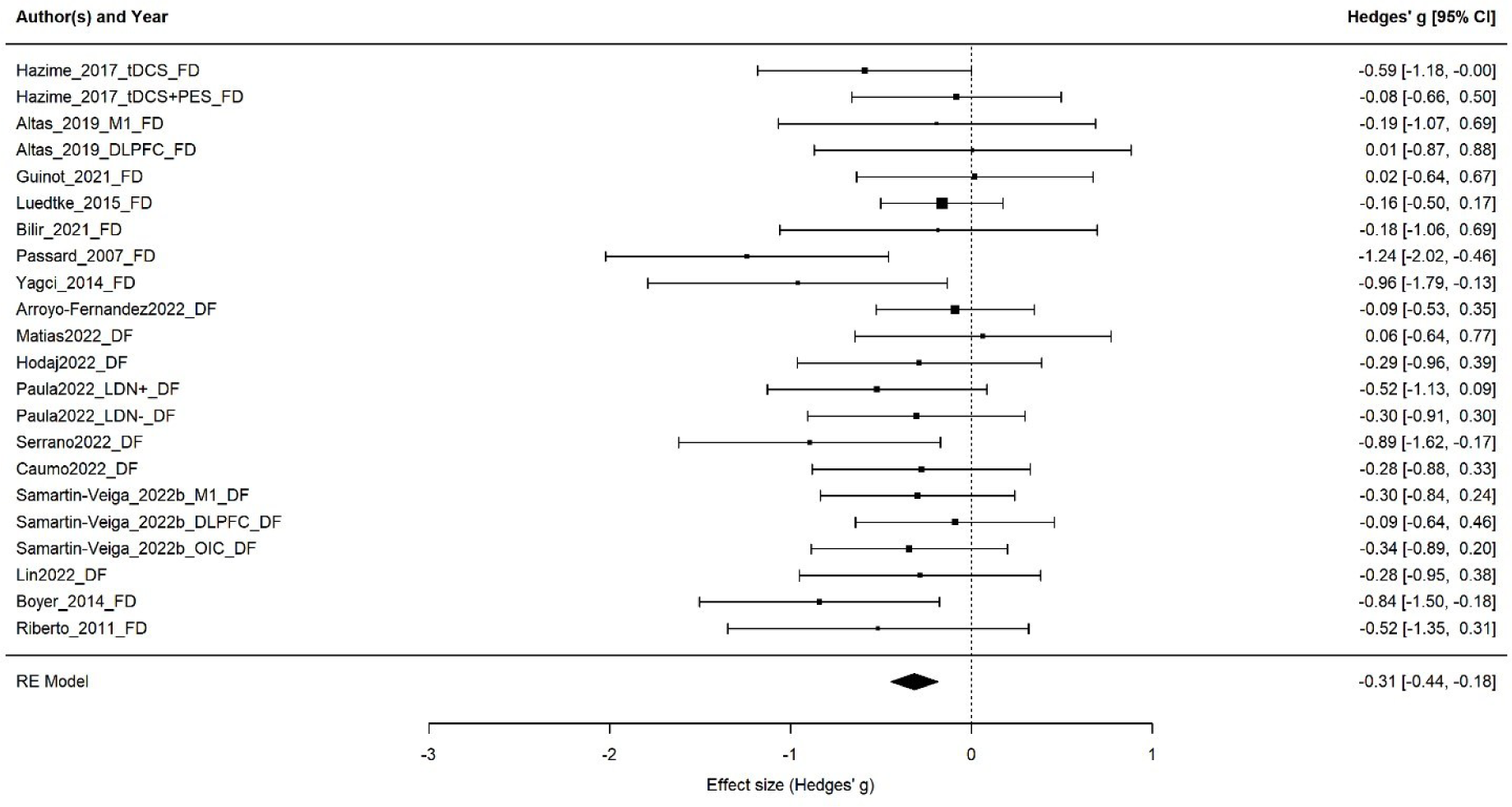
Forest plot of the effect size of noninvasive brain stimulation on functional disability change score. CI = confidence interval.

**Figure 8.**
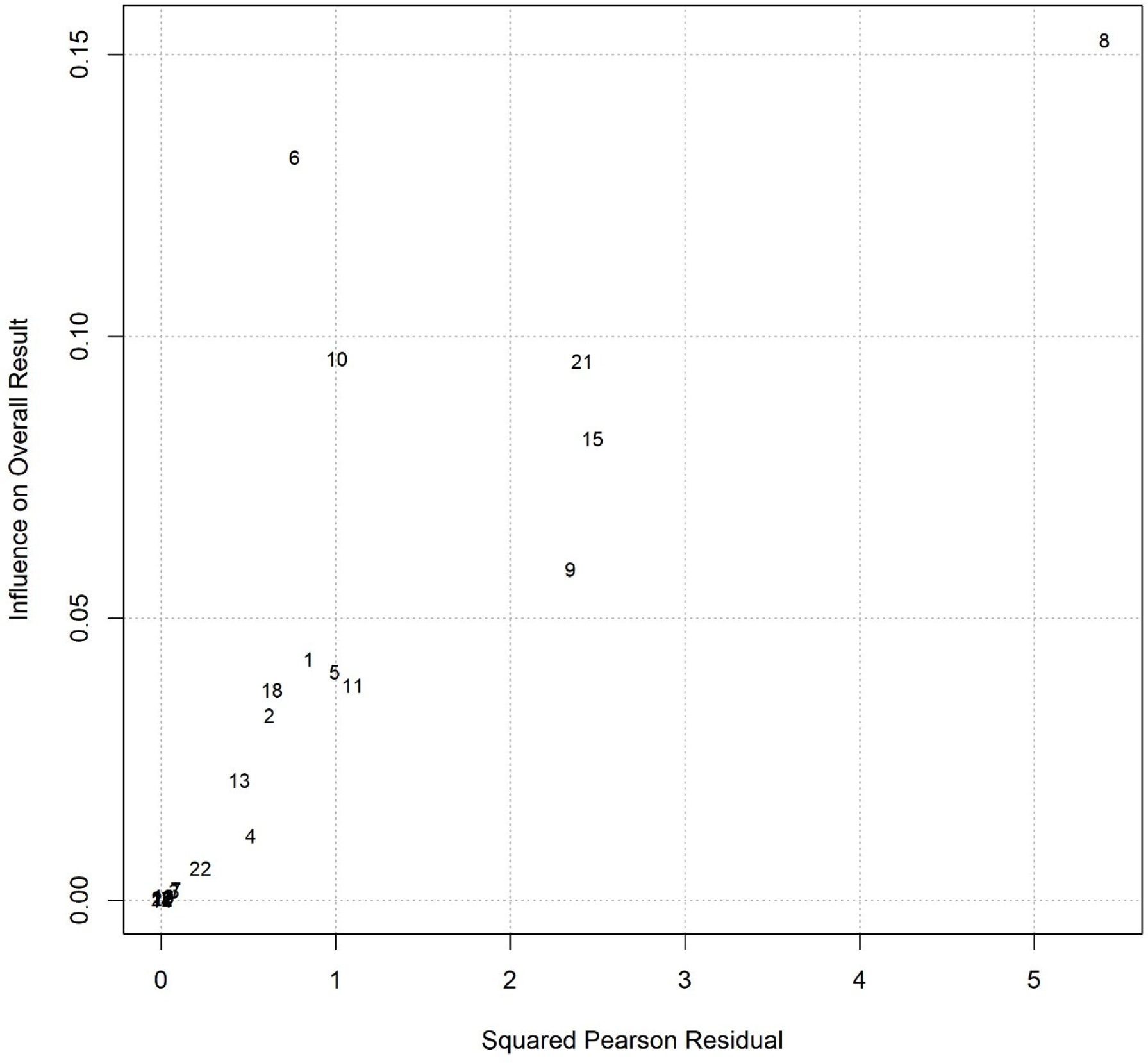
Baujat plot of studies distribution considering pre-post functional disability.

The included moderators and covariates did not highlight significant patterns (**Table S10** and **Table S11**).

Considering the publication bias, the Egger test modified version using the Pustejovsky-Rodgers’ method showed no asymmetry b = 0.35, 95% CI [-1.66, 2.36], which is not significantly different from zero, z = - 0.68, p = .494.

## Discussion

The CPP diagnosis has been included in the latest ICD revision (ICD-11) [170] based on the central sensitization hypothesis [179]. This theory posits that the central nervous system plays a critical role in modulating pain processing and experience, leading to hypersensitivity. Emerging evidence supports this theory, showing common neural underpinnings across chronic pain conditions now included in the CPP [10; 116]. This observation highlights the potential of NIBS to modulate maladaptive brain activity and connectivity to promote plastic changes [29; 92; 118; 180; 182].

Previous meta-analyses and reviews have explored the effectiveness of NIBS in reducing pain in specific disorders now included in CPP [21; 48; 70] or in chronic pain conditions regardless of their classification as primary or secondary conditions [1; 121]. However, to our knowledge, no previous study investigated the effects of NIBS on CPP as a unified category. To address this gap, we systematically reviewed and quantitatively analyzed the impact of NIBS on core CPP symptoms, namely pain severity, emotional distress, and functional disability in the short and medium term. We included thirty-five sham-controlled RCTs, comprising a total of 874 participants receiving the real stimulation and 713 receiving a sham one. The studies primarily included rTMS and tDCS protocols, with only one trial applying single-pulse TMS [13] or tACS [167] protocols. In line with the literature, studies typically selected target regions to be stimulated by the M1 and left DLPFC [38; 156]. Detailed participant and protocol characteristics are summarized in the results section and supplementary materials.

### NIBS short-term effects

Regarding the NIBS short-term effects observed immediately after the end of the treatment, we found that real stimulation significantly reduced pain severity scores compared to the placebo condition. This result aligns with previous meta-analyses that explored the effects of NIBS on migraine [48] and its role as an add-on therapy to drugs in fibromyalgia [70].

Real stimulation also showed a moderate, significant effect in improving functional disability scores. Functional disability refers to a person’s ability to carry out daily activities, such as walking and dressing, or participating in social or work-related roles [114].

Interestingly, previous meta-analyses or reviews evaluating the effects of NIBS on this outcome measure either did not report significant improvements [121; 131] or did not explore it altogether [21; 48]. It is worth noting that the meta-analysis by O’Connell et al. (2018) differs from the current work in two significant aspects related to the studies’ inclusion criteria. First, they did not disentangle between primary and secondary chronic pain conditions, and second, they excluded studies targeting migraine and other headache conditions.

We further considered quality of life as an outcome of interest, that is a broader and more subjective measure that includes physical functionality and emotional, psychological, and social well-being [26; 47], thus reflecting the overall level of satisfaction individuals feel in their lives. Concerning the quality of life, our findings did not show any significant effect of real stimulation compared to sham. We did not find this result surprising since the quality of life is a complex construct, and we did not expect a ‘direct’ effect of stimulation in this outcome. For the sake of clarity, it should also be acknowledged that only a limited number of the included studies (seven considering the pre-post intervention) measured quality of life; therefore, analyses on this variable should be considered preliminary.

Finally, a small but significant improvement in anxiety and depressive symptoms was observed in patients receiving real stimulation. Prior data synthesis works highlighted that NIBS can impact the affective domain in CPP, reporting reductions in depressive symptoms and anxiety [70] [21]. However, previous studies suggested that effects on pain severity and affective symptoms might depend on the targeted brain region. Specifically, Hou et al. (2016) and Brighina et al. (2019) reported that the stimulation of M1 was more effective in reducing pain intensity and fatigue while targeting the left DLPFC improved depressive and anxiety symptoms. Our findings did not highlight an effect of the target region in predicting pain intensity or emotional distress scores. However, it should be noted that our dataset was unbalanced in this sense, including more studies targeting the left M1 than the left DLPFC, thus preventing the emergence of differences between the stimulated target regions.

Overall, our findings on short-term effects suggest that NIBS is effective in modulating the core symptoms of CPP. While the exact neural mechanisms behind these improvements are not fully understood, it is known that the DLPFC and M1 are extensively connected to cortical and subcortical brain structures [83; 103] which are involved in multiple functions, from the regulation of cognitive and affective processes to the release of neurotransmitter systems that can ultimately affect the processing and experience of pain [38; 156]. Therefore, the stimulation produces effects at the local and distal levels, inducing large-scale network modifications based on anatomical and functional connectivity [57; 80; 94; 139; 149; 152; 159]. For instance, previous studies suggested neurotransmitters (e.g., glutamate, GABA, and serotonin) [17; 97; 110] and endogenous opioids release following M1 stimulation [38; 39; 99; 100]. The stimulation of DLPFC, on the other side, is known to modulate the activity of the anterior cingulate cortex [165; 166], which has been associated with emotional and pain processing [82]. Future studies should clarify the CPP pathological and healing neural mechanisms and whether one or more regions can be the optimal candidates for effective and sustained intervention in CPP.

Considering the other categorical moderators, our data did not highlight differences between the choice of rTMS and tDCS protocols, which aligns with the most recently updated expert panels’ guidelines for the clinical use of NIBS in chronic pain [51; 95] recommending a Level B ‘probably efficacy’ for tDCS and a Level A ‘definite efficacy’ for rTMS analgesic effects on neuropathic pain. Moreover, no differences emerged between improvements in anxiety and depressive symptoms, suggesting that the improvements were not driven by one of the two measures. The similar response of anxiety and depressive symptoms is not surprising since they are highly correlated [28; 75] and benefit similarly from NIBS intervention [27; 30; 175], possibly sharing some neural underpinnings [101].

We also explored the impact of continuous predictors, namely the number of pulses per session (only for pain intensity), the number of sessions, and illness duration. A trend toward significance was found for the illness duration on emotional distress change score. Specifically, a reduction of improvement in mood and anxiety symptoms was observed for real stimulation when patients suffered pain for longer, suggesting that longer illness duration predicted reduced benefits from the stimulation. This point will be addressed more thoroughly in the following paragraph.

### NIBS medium-term effects

As a secondary outcome measure, we included the one-month follow-ups measuring the effects of the three core symptoms of CPP [114]. Our findings highlight that the effect of real stimulation improving pain severity and functional disability remained significant one month after the end of the treatment. Conversely, no difference between sham and real stimulations was traceable on emotional distress.

Previous meta-analyses, including CPP patients, did not quantify the effects of NIBS on follow-up measures [48; 70]. O’Connell et al. (2018) [121] reported an effect of tDCS in follow-up measures comprised between one and six weeks post-treatment, whereas no clear evidence emerged for rTMS. The sustained effects of NIBS found at the one-month follow-up point to the involvement of metaplastic changes induced by the stimulation [71]. Meta-regression analyses also reinforced this hypothesis, suggesting an effect of illness duration and number of sessions in predicting stimulation outcomes. Specifically, illness duration significantly predicted the effectiveness of stimulation on pain intensity and emotional distress scores, with longer illness duration reducing the improvements induced by the stimulation. The number of sessions showed a trend toward significance in predicting the pain intensity, namely increasing the number of sessions predicted a higher reduction in symptoms. Such findings are in line with previous evidence. On one side, the impact of the number of sessions has been reported in the treatment of major depressive disorders [72; 154], and craving and consumption behaviors in eating and drug disorders [158] highlighting an improvement in outcome measures due to the cumulative effects of stimulation [147]. On the other side, a longer duration of (untreated) illness negatively affects treatment outcomes across several disorders, including schizophrenia [4; 134], eating disorders [12], and obsessive-compulsive disorders [135]. Such an effect has also been reported in studies including CPP participants, specifically chronic low back pain [171] and fibromyalgia [46]. The link between longer illness duration and more negative treatment outcomes can be based on neuroplastic maladaptive changes [53] that could possibly be more resistant to the treatment and, specific to our case, by NIBS. Of course, maladaptive neuroplastic changes are not the only factor explaining negative treatment outcomes; indeed, other mechanisms, including dysfunctional coping strategies such as catastrophizing thinking, rumination and helplessness feelings [58; 91] and cognitive impairment, such as attentional deficits, reduced working memory and hypervigilance or fear [8; 37; 109; 112; 136], may take place in chronic pain disorders [44; 55; 91; 106]. Observational studies showed that the observed avoidance behaviors or catastrophizing thinking, frequently reported in chronic pain patients, may exacerbate the experience of pain, the impact of the disease on individuals’ lives, and the maintenance of negative emotions and depression symptoms [40; 102; 163].

Therefore, combined interventions are required to consider the disorder’s multifaceted nature. In line with this evidence, literature is now moving towards the idea of multimodal interventions [153; 160; 175], in which the effects of NIBS may be maximized by combining the stimulation with behavioral or cognitive interventions. Cognitive/behavioral interventions may promote plastic changes at the brain level, addressing maladaptive coping strategies or dysfunctional processes through, for instance, psychotherapy interventions, such as cognitive-behavioral approaches, including acceptance and commitment therapy (NICE guidelines, 2021) [113], and such psychological changes can be further reinforced and sustained through NIBS induced plasticity.

In the current work, we tried to explore the effects of NIBS applied as monotherapies, add-ons, or augmentative interventions in the included studies. However, unfortunately, we could not quantify such effects due to the unbalanced samples of included articles. Indeed, most of the studies applied an add-on strategy, combining NIBS with patients’ ongoing pharmacotherapy, with only 4 studies applying NIBS as monotherapies and 8 using an augmentative strategy (Table 2 and 3). Moreover, research applying an augmentative strategy comprised very different approaches, combining stimulation with physical activity [11; 105; 125], mirror box [155], or multiple interventions, including relaxation and cognitive-behavioral therapy [60; 148]. Future studies are therefore necessary to clarify this point.

Considering the null results in emotional distress, it should be acknowledged that such an effect was small also immediately after the treatment. It is possible that the higher percentage of studies stimulating M1 in studies included in the pre-post analysis, which became the totality of studies included at the follow-up, contributed to such an effect.

To conclude, we ran correlations among the effect sizes of the four measures to investigate whether positive changes in one of the dimensions could be correlated to improvements in the others. No correlation was traceable among the outcome measures’ change scores despite NIBS being independently effective on the three core symptoms of CPP, namely pain intensity, emotional distress, and functional disability. It suggests that improvements in the three variables are based on at least partially different mechanisms. Considering follow-up measures, the three variables correlated each other, suggesting a possible stabilization or consolidation of effects due to individuals’ experiences in everyday lives.

### Study limitations

The current work presents several limitations. The first certainly relates to the heterogeneity in the stimulation protocols of the included studies, which was observed at qualitative and quantitative levels. Articles differed in the type of stimulation delivered, target regions, number and frequency of sessions, and stimulation parameters. A striking example, in this sense, concerns the side of stimulation for M1, which was left in the 48.5% of the cases [5; 11; 20; 34; 45; 68; 85; 96; 98; 105; 107; 108; 129; 148; 151; 162; 181], contralateral to the pain-affected area of the body in the 14.3% [62; 79; 125; 155; 177], ipsilateral [60] or contralateral [35] to the dominant district 2.8% and 2.8% respectively. The studies’ heterogeneity, reduced number of levels included in some of the analyses, and the few studies reaching a low-level risk of bias prevent the possibility of tracing clear operational guidelines considering how deliver NIBS in CPP disorders.

Second, most of the included studies relate to patients with chronic migraine or fibromyalgia, while only 9 of the included studies refer to other CPP conditions. Future studies are needed to clarify whether NIBS can effectively improve symptoms across specific CPP conditions.

Third, anxiety and depressive symptoms have been analyzed as emotional distress measures. This was done since most of the studies included these questionnaires, but it is important to note that they represent only some possible manifestations of emotional distress.

### Conclusions

In the current work, we systematically reviewed and quantitatively analyzed the effect of NIBS on CPP core symptoms, namely pain intensity, emotional distress, and functional disability.

Overall, our findings suggest that NIBS can effectively reduce the core symptoms of CPP in the short and medium term, highlighting an effect – in the opposite direction - of the number of stimulation sessions and illness duration. However, it suggests that NIBS alone may be insufficient for sustained improvement, especially in patients with long-standing CPP. The findings recommend a multimodal approach that combines NIBS with cognitive-behavioral therapies to address the complex neural and psychological components of chronic pain. Future research should investigate the optimal integration of NIBS with behavioral interventions and explore targeted protocols based on individual symptom profiles and illness history. Definitively, more studies are required to clarify CPP’s pathological and healing mechanisms to develop effective, evidence-based, and sustained changes.

## Data Availability

All data produced in the present study are available upon reasonable request to the authors

## Footnotes

1 In line with previous literature, we refer to ‘anodal’ tDCS for studies positioning the anode over the relevant region stimulated to modulate symptoms (and therefore using an excitatory strategy), to ‘cathodal’ tDCS when the cathode was positioned over the target area (thus applying an inhibitory strategy), or bi-hemispheric when the stimulation strategy consisted in the synchronous and polarity-opposite stimulation of two brain regions.

