## Supplemental Materials for "Non-Invasive Brain Stimulation for Core Symptoms of Chronic Primary Pain: A Systematic Review and Meta-Analysis of randomized controlled trials"

**Supplementary materials**

***Section A – Methods section details***

**Table S1**. Literature search strategy

| Database | Search strings |
|---|---|
| Pubmed (Jan 2023) | (("atypical facial pain"[Title/Abstract] OR "burning mouth syndrome"[Title/Abstract] OR "burning mouth syndrome"[MeSH Terms] OR "migraine disorders"[Title/Abstract] OR "migraine disorders"[MeSH Terms] OR "abdominal pain"[Title/Abstract] OR "abdominal pain"[MeSH Terms] OR "neck pain"[Title/Abstract] OR "neck pain"[MeSH Terms] OR "chest pain"[Title/Abstract] OR "chest pain"[MeSH Terms] OR "phantom limb"[Title/Abstract] OR "phantom limb"[MeSH Terms] OR "low back pain"[Title/Abstract] OR "low back pain"[MeSH Terms] OR "facial pain"[Title/Abstract] OR "facial pain"[MeSH Terms] OR "pelvic pain"[Title/Abstract] OR "pelvic pain"[MeSH Terms] OR "visceral pain"[Title/Abstract] OR "visceral pain"[MeSH Terms] OR "temporomandibular joint dysfunction syndrome"[Title/Abstract] OR "temporomandibular joint dysfunction syndrome"[MeSH Terms] OR "tension-type headache"[Title/Abstract] OR "tension-type headache"[MeSH Terms] OR "complex regional pain syndromes"[Title/Abstract] OR "complex regional pain syndromes"[MeSH Terms] OR "fibromyalgia"[Title/Abstract] OR "fibromyalgia"[MeSH Terms] OR "irritable bowel syndrome"[MeSH Terms] OR "irritable bowel syndrome"[Title/Abstract] OR "persistent idiopathic facial pain"[Title/Abstract] OR "small-fiber neuropathy"[Title/Abstract] OR "small-fiber neuropathy"[MeSH Terms] OR "trigeminal autonomic cephalalgias"[Title/Abstract] OR "trigeminal autonomic cephalalgias"[MeSH Terms] OR "trigeminal neuralgia"[Title/Abstract] OR "trigeminal neuralgia"[MeSH Terms] OR "interstitial cystitis"[Title/Abstract] OR "Hypertrophic Gastritis"[Title/Abstract] OR "Hypertrophic Gastritis"[MeSH Terms] OR "Prostatitis"[Title/Abstract] OR "Prostatitis"[MeSH Terms] OR "idiopathic pain"[Title/Abstract] OR "primary pain"[Title/Abstract])) AND ("transcranial magnetic stimulation"[MeSH Terms] OR "transcranial magnetic stimulation"[Title/Abstract] OR "tms"[Title/Abstract] OR "transcranial direct current stimulation"[Title/Abstract] OR "transcranial direct current stimulation"[MeSH Terms] OR "tdcs"[Title/Abstract]) |
| Scopus (Jan 2023) | (TITLE-ABS-KEY ( "atypical facial pain" ) OR TITLE-ABS-KEY ( "chronic burning mouth" ) OR TITLE-ABS-KEY ( "chronic migraine" ) OR TITLE-ABS-KEY ( "chronic primary abdominal pain syndrome" ) OR TITLE-ABS-KEY ( "chronic primary bladder pain syndrome" ) OR TITLE-ABS-KEY ( "chronic primary cervical pain" ) OR TITLE-ABS-KEY ( "chronic primary chest pain syndrome" ) OR TITLE-ABS-KEY ( "chronic primary epigastric pain syndrome" ) OR TITLE-ABS-KEY ( "Chronic primary limb pain" ) OR TITLE-ABS-KEY ( "chronic primary low back pain" ) OR TITLE-ABS-KEY ( "chronic primary orofacial pain" ) OR TITLE-ABS-KEY ( "chronic primary pelvic pain syndrome" ) OR TITLE-ABS-KEY ( "chronic tension-type headache" ) OR TITLE-ABS-KEY ("chronic temporomandibular disorder pains") OR TITLE-ABS-KEY ( "complex regional pain syndromes" ) OR TITLE-ABS-KEY ( "fibromyalgia" ) OR TITLE-ABS-KEY ( "irritable bowel syndrome" ) OR TITLE-ABS-KEY ( "persistent idiopathic facial pain" ) OR TITLE-ABS-KEY ( "small-fiber neuropathy" ) OR TITLE-ABS-KEY ( "trigeminal autonomic cephalalgias" ) OR TITLE-ABS-KEY ( "trigeminal neuralgia" ) OR TITLE-ABS-KEY ( "chronic primary thoracic pain" ) OR TITLE-ABS-KEY ( "chronic primary visceral pain" ) OR TITLE-ABS-KEY ( "interstitial cystitis" ) OR TITLE-ABS-KEY ( "idiopathic pain" ) OR TITLE-ABS-KEY ( "primary pain" )) AND (TITLE-ABS-KEY ( "transcranial magnetic stimulation" ) OR TITLE-ABS-KEY ( "tms" ) OR TITLE-ABS-KEY ( "transcranial direct current stimulation" ) OR TITLE-ABS-KEY ( "tdcs" ) OR TITLE-ABS-KEY ( "repetitive transcranial magnetic stimulation" ) OR TITLE-ABS-KEY ( "rtms" )) |
| Embase (Jan 2023) | ('atypical facial pain'/exp OR 'atypical facial pain' OR 'burning mouth syndrome':ti,ab,kw OR 'migraine'/exp OR 'migraine' OR 'abdominal pain'/exp OR 'abdominal pain' OR 'neck pain':ti,ab,kw OR |

| | |
|---|---|
| | 'chest pain':ti,ab,kw OR 'phantom limb'/exp OR 'phantom limb' OR 'low back pain':ti,ab,kw OR 'face pain'/exp OR 'face pain' OR 'pelvic pain'/exp OR 'pelvic pain' OR 'visceral pain'/exp OR 'visceral pain' OR 'temporomandibular joint disorder'/exp OR 'temporomandibular joint disorder' OR 'tension headache'/exp OR 'tension headache' OR 'complex regional pain syndrome'/exp OR 'complex regional pain syndrome' OR 'fibromyalgia'/exp OR 'fibromyalgia' OR 'irritable bowel syndrome':ti,ab,kw OR 'persistent idiopathic facial pain'/exp OR 'persistent idiopathic facial pain' OR 'small-fiber neuropathy'/exp OR 'small-fiber neuropathy' OR 'trigeminal autonomic cephalalgia'/exp OR 'trigeminal autonomic cephalalgia' OR 'trigeminal neuralgia':ti,ab,kw OR 'interstitial cystitis'/exp OR 'interstitial cystitis' OR 'menetrier disease'/exp OR 'menetrier disease' OR 'prostatitis'/exp OR 'prostatitis' OR 'idiopathic pain'/exp OR 'idiopathic pain' OR 'primary pain':ti,ab,kw) AND ('transcranial magnetic stimulation'/exp OR 'transcranial magnetic stimulation' OR 'tms'/exp OR 'tms' OR 'transcranial direct current stimulation'/exp OR 'transcranial direct current stimulation' OR 'tdcs') |
| Web of Science (Jan 2023) | TS=('atypical facial pain' OR 'chronic burning mouth' OR 'chronic migraine' OR 'chronic primary abdominal pain syndrome' OR 'chronic primary bladder pain syndrome' OR 'chronic primary cervical pain' OR 'chronic primary chest pain syndrome' OR 'chronic primary epigastric pain syndrome' OR 'chronic primary limb pain' OR 'chronic primary low back pain' OR 'chronic primary orofacial pain' OR 'chronic primary pelvic pain syndrome' OR 'chronic tension-type headache' OR 'chronic temporomandibular disorder pains' OR 'complex regional pain syndromes' OR 'fibromyalgia' OR 'irritable bowel syndrome' OR 'persistent idiopathic facial pain' OR 'small-fiber neuropathy' OR 'trigeminal autonomic cephalalgias' OR 'trigeminal neuralgia' OR 'chronic primary thoracic pain' OR 'chronic primary visceral pain' OR 'interstitial cystitis' OR 'idiopathic pain' OR 'primary pain') AND TS=('transcranial magnetic stimulation' OR 'tms' OR 'repetitive transcranial magnetic stimulation' OR 'rtms' OR 'transcranial direct current stimulation' OR 'tdcs') |

| **Table S2:** Additionally participants' characteristics from the included studies. | |
|---|---|
| **TMS protocols** | |
| **Authors** | **Place of recruitment** |
| Ali AbdElkader et al., 2021 | Out-Patient Clinic |
| Altas et al., 2019 | Physical Medicine And Rehabilitation |
| Avery et al., 2015 | Advertising, Publicity On Television, And A Specialized Hospital Clinic |
| Bilir et al., 2021 | Physical Medicine And Rehabilitation Outpatient Clinics |
| Boyer et al. 2014 | La Timone University Hospital (Marseille, France) |
| Guinot et al., 2021 | Pain And Rheumatology Department |
| Mhalla et al. 2011 | N.R. |
| Misra et al., 2013 | Out-Patient Clinic |
| Passard et al., 2007 | N.R. |
| Picarelli et al., 2010 | N.R. |
| Tekin et al., 2014 | Sisli Etfal Education And Research Hospital Physical Medicine And Rehabilitation Outpatient Unit |
| Todorov et al., 2020 | N.R. |
| Umezaki et al. 2016 | Medical University Of South Carolina + Emails |
| YAĞCI et al., 2014 | N.R. |
| **Tes Protocols** | |
| **Authors** | **Place Of Recruitment** |
| Antal et al., 2011 | N.R. |
| Arroyo-Fernandez, 2022 | FM Patient Associations |
| Caumo, 2022 | Hospital De Clınicas De Porto Alegre (Brazil) |
| de Melo et al., 2020 | N.R. |
| Dutra et al., 2020 | Natal (Brazil) |
| Fagerlund et al., 2015 | Pain Clinic, University Hospital Of Northern Norway, Tromsø |
| Hazime et al., 2017 | University Of Sao Paulo) And Rehabilitation Centre (Irmandade Santa Casa De Misericordia Of Sao Paulo) |
| Hodaj et al.,2022 | Pain Centre Of The Grenoble Alpes University Hospital |
| Jiang et al. 2020 | University Hospital |
| Khedr et al., 2017 | Assiut University Hospital |
| Lin, 2022 | Taipei Medical University Hospi- Tal. |
| Luedtke et al., 2015 | N.R. |
| Matias, 2022 | Federal University Of Rio Grande Do Norte |
| Oliveira et al. 2015 | Adventist College Of Bahia, Brazil. |

| | |
|---|---|
| Paula, 2022 | La Salle Saude, Canoasn.R.RS, Brazil. |
| Riberto et al. 2011 | N.R. |
| Samartin-Veiga, 2022 | Patients Included In Previous Research, Local Health Centers, Press, Patient Associations |
| Segal, 2021 | Lowenstein Rehabilitation Hospital (Ra'anana, Israel) |
| Serrano, 2022 | Hospital De Clinicas De Porto Alegre (HCPA) |
| To et al. 2017 | The University Hospital Antwerp, Belgium |
| Volz et al., 2016 | Gastroenterology, Infectious Diseases, Rheumatology |
| Notes: n.r. = not reported | |

*Study quality assessment*

Two authors (A. T., A. Vedani) performed quality assessment independently using the Cochrane Collaboration's Risk-of-Bias Tool for RCT [39] which focuses on five domains: *1) Bias arising from the randomization process,* assessing whether the allocation sequence was properly generated and concealed to prevent selection bias; *2) Bias due to deviations from intended interventions*, which evaluates whether participants and personnel were blinded to the interventions and whether any deviations from the intended interventions occurred. Studies were evaluated as low-risk whether it *; 3) Bias due to missing outcome data*, assessing the completeness of outcome data and whether any missing data could affect the results; 4) *Bias in the measurement of the outcomes,* evaluating how outcomes were measured and whether knowledge of the intervention by outcome assessors could have influenced the results; 5) *Bias in the selection of the reported result*, assessing whether outcomes were selectively reported, such as including only positive findings or omitting non-significant results. Conflicts were solved by consensus of the two researchers and a third researcher was consulted when needed (A.Vergallito).

*Outcomes measures' details*

Studies varied in the outcome measures used, typically including self-administered questionnaires to assess pain severity, emotional distress, functional impact of the disease, and quality of life.

*Pain severity* typically included the Visual Analogue and Number Rating Scale (VAS and NRS) ratings. Papers reporting only a measure of pain frequency to indicate pain severity [20; 26] were not included in the analysis. Indeed, frequency is typically considered a measure of pain only for migraines but not other CPP conditions. Effects of NIBS on migraine attack frequency can be found in other meta-analyses [8; 26; 36].

E*motional distress* was measured through clinicians-administered or self-report standardized questionnaires investigating anxiety and depressive symptoms, such as the Hospital Anxiety and Depression Scale (HADS) [38], the Beck Anxiety Inventory (BAI)[18], the Beck Depression Inventory (BDI) [2], the Hamilton Depression Rating Scale (HDMS) [14] and the Hamilton Anxiety Rating Scale (HARS) [13], the Montgomery Asberg Rating Scale (MADRS) [23], and the State-Trait Anxiety Inventory (STAI) [37].

F*unctional disability* was investigated through the Roland-Morris Disability Questionnaire (RMDQ) [33], or the Oswestry Disability Index (ODI) [33].

As secondary endpoints, we considered the pre-post scores of *quality of life*, assessed through standardized questionnaires such as the Short Form Health Survey 36 (SF-36) [46], or the World Health Organization Quality-of-Life Scale – Brief version (WHOQOL-BRIEF)[43], and the follow-up assessment of each outcome measure (Table S3).

It is crucial to highlight that since the IASP definition of CPP is very recent, the papers included were heterogeneous in including or not the previous outcome measures.

Considering the aims of our work, one of the authors (AT) categorized the reported scales into the three core symptoms according to the construct measured by each questionnaire.

*Quantitative analyses*

We extracted relevant information for each article, comprising NIBS protocol features (specific technique, session number, target regions), the number of patients included in experimental and control groups and their demographic and clinical features, means and standard deviations from pre-, post-treatment, and follow-up outcome measures. We contacted the authors to obtain the missing data when we found insufficient information in the paper's text, tables, or supplementary materials. When data were available in graphical presentations, we used the free software WebPlotDigitizer (https://automeris.io/WebPlotDigitizer/) to extract them.

For the different outcome measures, we computed the pre-post-treatment mean difference (subtracting pre-treatment measures from post-treatment measures so that a negative value represents symptom improvement) for the experimental and the control groups. We calculated the standard deviation of the pre-post treatment difference according to the Cochrane Handbook for systematic reviews of intervention guidelines[5]:

$$SDchange = \sqrt{SDpre^2 + SDpost^2 - (2 * corr * SDpre * SDpost)}$$

Where *corr* represented the correlation between pre- and post-measurement variances and was set at .5 following Follman and colleagues[10].

We computed the sampling variance and standardized mean difference (SMD) for each included study using the *escalc* function of the metafor package for R, version 3.4.3 [45]. The SMD function automatically corrects for the positive bias due to small groups [17; 45], computing the Hedge's *g*, used in the present work as an effect size measure.

Considering the included studies, some had sufficient information to calculate more than one effect size per outcome measure. For instance, considering pain intensity, a few papers included more than one group receiving NiBS over different target regions (i.e., Todorov 2020, Samartin Veiga 2022 [34; 41]). Similarly, emotional distress measures often included separate effect sizes of anxiety and depressive symptoms (i.e., [3; 7; 24] Fagerlundt 2015, Luedtke 2015; Bilir 2021). Considering these effect sizes as statistically independent would violate the independence assumption of traditional meta-analyses and bias the statistical findings. Therefore, to address this issue, we ran a multi-level random effects model using the *rma.mv* function of the metafor package [11], clustering the individual effect sizes at the study level. Then, in line with methodological guidelines [15], we compared the multi-level model with a reduced model (not including the three-level) using the *anova* function. We then applied the best-fitting model to analyze the data (see the Supplementary materials for details on results and procedures).

When no differences emerged between the two models, we accepted the simpler one [15]. In this case, we used a random-effects model with the *rma* function of the metafor package. We chose a random-effects model because it is suitable for dealing with heterogeneity due to sampling error and variance between studies' effect sizes [9].

We provided several measures to display data heterogeneity[15]. We reported the variation due to the sampling error (Q statistic), the percentage of variation between studies not linked to the sampling error ($I^2$ statistics) [19], and the prediction intervals (PIs) [21], an interesting measure that provides a range within which one can expect future studies effects to fall, based on the currently available data.

We planned to identify potential outliers and influential cases using the influence function *inf* implemented in the metafor package for the two-level analyses. For the multi-level models, potential outliers were identified through visual inspection and data plotting using the Cook distance measure [45]. As recommended by previous authors [45], when we detected extreme values, we removed them and refitted the model to verify that their elimination did not impact the analysis results. Then, we ran meta-regression analyses to investigate the effect of potential categorical and continuous variables that could explain the heterogeneity and magnitude of extreme values. Potentially interesting moderators were defined a priori and included the specific stimulation technique (tDCS, or rTMS), the target region (M1, DLPFC), the protocol type (inhibitory vs. excitatory), the specific disorder, and the blinding (single vs. double). The number of sessions, TMS pulses, and illness duration were hypothesized as possible continuous predictors. However, we could not include all the hypothesized factors, but we included only the informative ones — namely those that were sufficiently represented in the selected papers. As a rule of thumb, Cochrane guidelines suggest that subgroup analyses and meta-regressions should be run only when at least ten studies are available [4]. Considering the exploratory nature of moderators in our heterogeneous sample of studies, we performed subgroup analyses when at least six studies per group were available. Detailed meta-regressions are reported in the supplementary materials, whereas the main results are briefly discussed in the following paragraphs.

Considering publication bias analyses, guidelines suggest avoiding them when the between-study heterogeneity is high ($I^2 \approx 75\%$) since results are unreliable [15; 44]. When heterogeneity was lower, we used a modified version of the Egger regression test outlined by Pustejovsky and Rodgers (2019)[29] (see Harrer et al., 2021[15]). Indeed, for SMD effect sizes, the Egger regression test can inflate false positive results due to the non-independence of standardized mean differences and standard errors. Pearson's correlations were run to explore correlations among symptom changes after the treatment and at follow-up using the Hmisc package and plotted using the Corrplot package [47].

#### *Section B – Qualitative synthesis of the included RCTs*

##### *Overall outcome measures*

In this systematic review, the effects of NIBS in CPP were measured on the three core symptoms of the disease [27]: pain intensity, emotional distress, and functional disability. Since some articles included it, we also explored NIBS's effects on the participants' reported quality of life.

It should be considered that CPP diagnosis is very recent; therefore, while pain intensity was the primary outcome for most of the studies included, not all of them addressed emotional distress and functional disability. Moreover, none of the included studies directly addressed CPP patients; instead, they targeted specific chronic pain conditions now included in CPP. Keeping this in mind, of the 14 studies applying TMS, all assessed pain intensity, 11 (73%) additionally included measures of emotional distress, and 9 (60%) also assessed functional disability. Among the 20 studies applying tDCS and one applying tACS, all but one study (95%) assessed pain intensity, 14 (60%) assessed emotional distress, and 13 (56%) measured functional disability. As previously reported, only five studies per group (33% of TMS studies; 21% of tDCS studies) also included a quality-of-life measure (Figure S1 graphically summarizes the outcome measures distribution).

**Table S3**: Additionally qualitative data on TMS protocols used in the included studies

| Study | Coil position | Sham procedure | Side effects | Medication |
|---|---|---|---|---|
| **Ali AbdElkader et al., 2021** | tangentially over the left DLPFC cortex | Coil placed at 90° to the targeted area | N.r. | N.r. |
| **Altas et al., 2019** | DLPFC: F3; vertex: CZ<br>M1: motor hotspot | Reverse positioned coil over vertex (0.1 Hz. %1 RMT) | N.r. | Stable treatment for 4 weeks |
| **Avery et al., 2015** | Left forehead | Sham coil | N.r. | Allowed |
| **Bilir et al., 2021** | Left DLPFC | Reverse positioned coil | No adverse events reported | Stable treatment for 12 weeks |

| | | | | |
|---|---|---|---|---|
| **Boyer et al. 2014** | Left M1 | Sham coil | No side effects | Stable treatment for 4 weeks |
| **Guinot et al., 2021** | M1 cortex (dominant thenar area) | Sham coil | N.r. | Stable treatment for 12 weeks |
| **Mhalla et al. 2011** | Left M1 | Sham coil | Headache, headache lasting <3 hours, dizziness | Stable treatment for 4 weeks |
| **Misra et al., 2013** | on the left PFC corresponding to the hot spot of the right | Sham coil | drowsiness for 12 h | Stable treatment for 4 weeks |
| **Passard et al., 2007** | Left M1 | Sham coil | N.r. | Stable treatment for 4 weeks |
| **Picarelli et al., 2010** | M1 contralateral to the painful upper limb | Sham coil | Headache, Neck pain, Dizziness. generalized seizure, after the seventh rTMS session | Stable treatment for 12 weeks |
| **Tekin et al., 2014** | M1 | Sham coil | Mild and transient headache | No medications |
| **Todorov et al., 2020** | M1 and left DLPFC | In Coil placed at 90° to the targeted area | N.r. | N.r. |
| **Umezaki et al., 2016** | F3 | Sham coil | Headache | Stable treatment for 4 weeks |
| **YAĞCI et al., 2014** | Left M1 | Coil placed at 90° to the targeted area | Transient headache, daily tinnitus | Stable treatment |

**Table S4.** Additionally qualitative data on tES protocols used in the included studies

| Study | Target region | Sham procedure | Side effects | Medication |
|---|---|---|---|---|
| **Antal et al., 2011** | Visual Cortex V1 | 30-second ramp up/ ramp down | mild tingling sensation headache, transient mild first-degree burnt, decreased appetite, rash, itching, headache with dizziness. | Ongoing treatment allowed |
| **Arroyo-Fernandez et al., 2022** | Left M1 | 30-second ramp up/ramp down | No severe adverse effects | Ongoing treatment allowed |
| **Caumo et al., 2022** | Left DLPFC | 30-second ramp up/ramp down | Burning sensation, headache, neck pain, mood swings, concentration difficulties | Ongoing treatment allowed |
| **de Melo et al., 2020** | Left M1 | 30-second ramp up/ramp down | No side effects reported | Ongoing treatment allowed |
| **Dutra et al., 2020** | Left DLPFC | 30-second ramp up/ramp down | skin tingling | n.r. |
| **Fagerlund et al., 2015** | Left M1 | 8-second fade-in period + 30 sec of direct current stimulation + 5-second fade-out | Headache, neck pain, scalp pain, tingling, itching, burning sensations, skin redness, sleepiness, trouble in concentration and acute mood change | Ongoing treatment allowed |
| **Hazime et al., 2017** | M1 contralateral to painful area | 30-second ramp up/ramp down | Headache, neck pain, scalp pain, back pain, tingling, itching, redness, burning sensations, sleepiness, trouble concentrating, nausea, mood change | Treatment allowed included analgesics, NSAIDs, opioids, antidepressants, muscle relaxants but no psychoactive drugs |
| **Hodaj, 2022** | Left M1 | 30-second ramp up/ramp down | No side effects reported | Ongoing treatment allowed |
| **Jiang et al. 2020** | M1 contralateral to painful/dominant area | 20-second ramp up/ramp down | itching, tingling, burning sensation, pain or warmth | Not allowed |
| **Khedr et al., 2017** | Left M1 | 30-second ramp up/ramp down | Itching, redness of skin | Itching and redness skin |
| **Lin et al., 2022** | Left M1 | 10-second ramp up/ramp down | Headache, neck pain, scalp pain, stinging, itch, burning sensation, drowsiness, concentration difficulties | Ongoing treatment allowed |

| | | | | |
|---|---|---|---|---|
| **Luedtke et al., 2015** | Left M1 | 30-second ramp up/ramp down | minimal transitory side effects | Ongoing treatment allowed |
| **Matias et al., 2022** | Left M1 | 30-second ramp up/ramp down | Headache, tingling, dizziness, nausea | Ongoing treatment allowed |
| **Oliveira et al., 2015** | M1 contralateral to painful area | 30-second ramp up/ramp down | No side effects | Ongoing treatment allowed |
| **Paula et al., 2022** | M1 contralateral to dominant area | 30-second ramp up | tingling, itching, and blushing. Headache, neck ache, scalp pain, burning sensation, sleepiness, and acute mood changes | Stable treatment for 12 weeks |
| **Riberto et al., 2011** | lM1 | 30-second ramp up/ ramp down | No side effects reported | Ongoing treatment allowed |
| **Samartin-Veiga et al., 2022** | Left M1 Left DLPFC Left OIC | 15-second ramp up/ ramp down | No side effects reported | Ongoing treatment allowed |
| **Segal et al., 2021** | M1 contralateral to painful area | 1-minute of ramping up to 1.5 mA and then back to 0 mA | Redness of the skin under the area of the electrode, itching sensation | Ongoing treatment allowed |
| **Serrano et al, 2022** | left DLPFC | 30-second ramp up/ramp down | mild side effects | Ongoing treatment allowed |
| **To et al., 2017** | C2 nerve dermatome and bilateral DLPFC | 5-second ramp up/ ramp down | No side effects reported | Ongoing treatment allowed |
| **Volz et al., 2016** | M1 contralateral to painful area | 30-second ramp up/ramp down | Headache, Scalp pain, Scalp burning sensation, Tingling, Itching, Skin redness, Sleepiness | Ongoing treatment allowed |

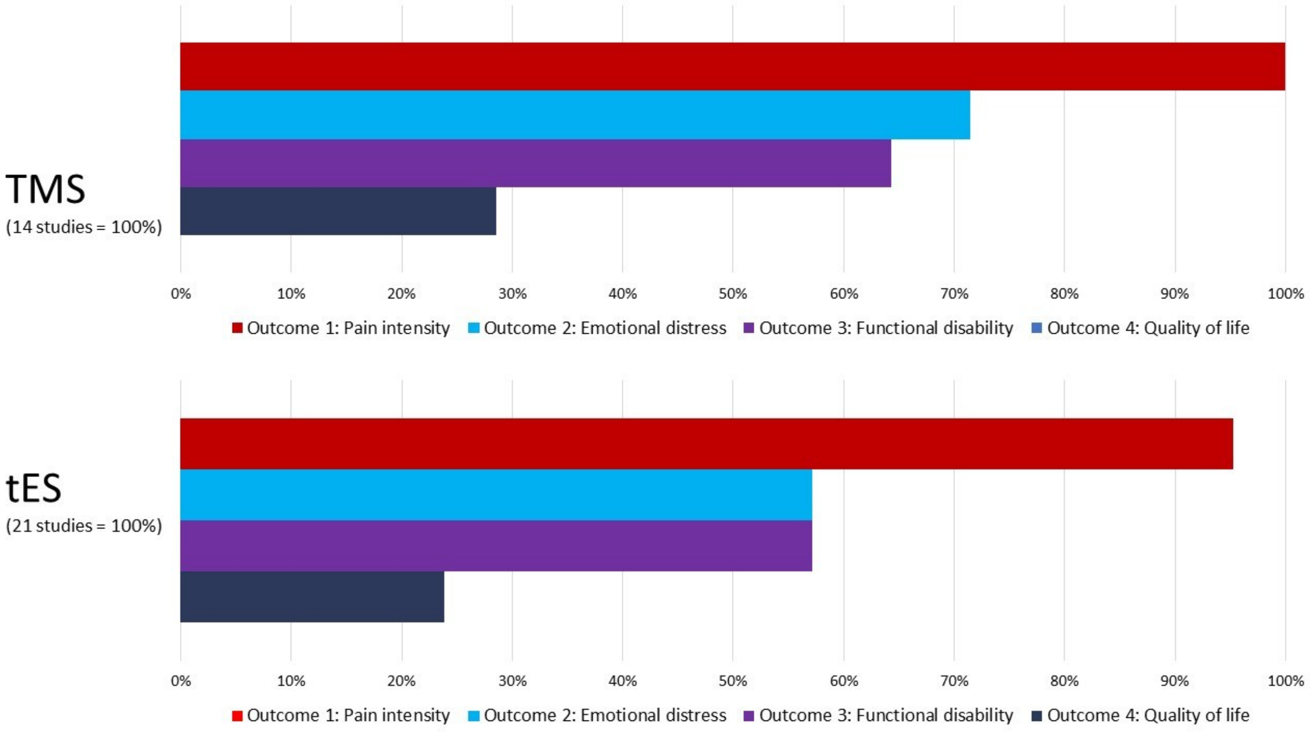


**Figure S1** shows the number of studies assessing certain outcomes' measures.

*Therapeutic strategies – details of studies applying augmentative and add-on NIBS techniques*

Following the definition of Razza and colleagues (2020)[31], we defined the treatment strategy as monotherapy when either TMS or tDCS are applied as the only treatment, i.e., patients were not treated with other approaches, neither were taking medication; "add-on" treatment strategy applies to studies where stimulation was added to the ongoing pharmacological therapy. In this case the two treatments, NIBS on one side and pharmacological therapy on the other side, were added and thought to work parallelly. Finally, "augmentative" treatment strategy refers to the therapeutic strategy in which TMS or tDCS are applied with other treatments, including experimental drugs. In this case, NIBS were not simply added but considered as an adjuvant approach to improving the effects of the other ongoing treatments.

In the study of Guinot and colleagues (2021), patients attended a 14-week multimodal training comprising an intensive phase, in which five daily rTMS sessions were delivered in the first two weeks, and a maintenance phase, in which participants receive two sessions of rTMS in the third week and one session from week 4 to week 14.

The stimulation sessions were combined with 3 hours of physical activity, including aerobic training, pool-based exercises, and relaxation three times per week [12]. Three studies added a program of physical exercises to the tDCS stimulation. The exercises included aerobic exercise sessions [1; 28], or warm-up (bicycle or walking), physical conditioning using neuromuscular (strength and localized muscular resistance and power) and neuromotor exercises (agility, coordination, and balance), and cool-down (self-stretching) [25].

Riberto and collaborators (2011) developed a complex rehabilitation program with a total duration of four months in which tDCS was added to psychoeducation or cognitive behavioral therapy groups focused on pain, physical and occupational therapy. In this program, tDCS was run before the first hour of activities of the rehabilitation program to test if it may act as a facilitator of behavioral modifications [32].

Segal and colleagues (2021) concurrently administered ten tDCS sessions with mirror therapy patients affected by phantom limb pain [35]. This rehabilitation technique uses the reflection of movements from a healthy limb in a mirror to create the illusion of the same movement in a paralyzed or impaired limb [30].

Considering the application of augmentation strategy combining stimulation and pharmacotherapy, de Paula et al. (2023) tested the combination of Naltrexone (real or placebo), an analgesic drug, and tDCS in fibromyalgia [6]. Finally, Hazime and colleagues (2017) combined real and sham peripheral electrical stimulation (PES) to tDCS to explore the top-down modulatory analgesic effects induced by central (tDCS) and peripheral (PES) neuromodulation techniques [16].

***Section B – Statistical Details and Meta-Regressions of Primary Outcome Measures***

This section highlights statistical details considering the primary outcome measures, from model selection to meta-regressions, including moderators (subgroup analyses) and predictors. We defined a-priori clinically relevant moderators and predictors, but we explored them only when they were represented in the studies included in the meta-analysis.

*Models' selection*

Model comparisons for the primary and secondary outcome measures are provided in Table S3. The model comparison was not performed for the quality-of-life assessment since no paper included more than one effect size.

**Table S5.** Models' selection.

| Model | **df** | **AIC** | **LRT** | **p (LRT)** |
|---|---|---|---|---|
| ***Primary endpoints: Pain intensity pre-post treatment*** | | | | |
| **Full model** | 3 | 76.66 | | |
| Reduced model | **2** | **75.90** | 1.24 | .266 |
| ***Primary endpoints: Emotional distress pre-post treatment*** | | | | |
| Full model | **3** | **35.82**# | | |
| **Reduced model** | 2 | 42.61 | 8.78 | .003 |
| ***Primary endpoints: Functional disability pre-post treatment*** | | | | |
| **Full model** | 3 | 18.04 | | |
| Reduced model | **2** | **16.04** | 0 | 1.00 |
| ***Secondary endpoints: Pain intensity pre to follow-up*** | | | | |
| **Full model** | 3 | 29.58 | | |
| Reduced model | **2** | **28.67** | 1.09 | .296 |
| ***Secondary endpoint: Emotional distress pre to follow-up*** | | | | |
| Full model | **3** | **18.78**# | | |
| **Reduced model** | 2 | 20.63 | 3.85 | .050 |
| ***Secondary endpoints: Functional disability pre to follow-up*** | | | | |
| **Full model** | 3 | 18.69 | | |
| Reduced model | **2** | **18.49** | 1.80 | .179 |

Table S5 summarizes the model selection procedure comparing the Full model, namely the three-level regression model, and the reduced model, which does not include the three-level. The best-fitting model is boldly highlighted.

### The multi-level model showed the best fit when comparing the emotional distress models. However, the analyses suggested that the variability was explained only by the cluster level, representing the between-studies heterogeneity variance computed in conventional two-level meta-analyses. Conversely, the variance explained by the nested level was 0. Therefore, following the previously cited guidelines (Harrer et al., 2021), the reduced model was applied to analyze the collected data.

*Pain intensity change score before and after the intervention*

**Table S6.** Subgroup analysis in pre-post pain intensity change score.

| Moderator / Subgroup analysis | g | SE | LL | UL | z | p | k | $p_{subgroup}$ |
|---|---|---|---|---|---|---|---|---|
| | | | | ***Pain intensity pre-post*** | | | | |
| ***NIBS type*** | | | | | | | | .359 |
| **tDCS** | -0.476 | 0.141 | -0.752 | -0.199 | -3.374 | < .001 | 20 | |
| **rTMS** | -0.710 | 0.213 | -1.128 | -0.292 | -3.329 | < .001 | 12 | |
| ***Target region*** | | | | | | | | > .123 |
| **lDLPFC** | -1.046 | 0.368 | -1.767 | -0.326 | -2.845 | .004 | 7 | |
| **lM1** | -0.328 | 0.160 | -0.642 | 0.014 | -2.044 | .041 | 14 | |
| **cpM1** | -0.821 | 0.229 | -1.271 | -0.371 | -3.577 | <.001 | 6 | |

Table S6 summarizes the categorical moderators and subgroup analyses. Note: g = Hedges' g effect size; SE = standard error of the coefficient; LL = lower limit of the 95% CI; UL = upper limit of the 95% CI; z = z-score associated with the g value in the same row; p = p-value associated with the z-score in the same row; k = the number of effect sizes contributing to g in the same row; psubgroup = p-value of the subgroup comparison.

**Table S7.** Covariates analysis in pre-post pain intensity change score.

| Covariate | Estimate | SE | LL | UL | z | p | k | Q | p | R2 (%) |
|---|---|---|---|---|---|---|---|---|---|---|
| **Illness duration** | | | | | | | 21 | 0.676 | .411 | 0 |
| **Intercept** | -0.836 | 0.266 | -1.358 | -0.314 | -3.137 | .002 | | | | |
| **Slope** | 0.026 | 0.032 | -0.036 | 0.088 | 0.822 | .411 | | | | |
| **Number of sessions** | | | | | | | 33 | 1.100 | .294 | 0 |
| **Intercept** | -0.797 | 0.275 | -1.336 | -0.257 | -2.894 | .004 | | | | |
| **Slope** | 0.025 | 0.024 | -0.022 | 0.073 | 1.049 | .294 | | | | |
| **Number of pulses** | | | | | | | 12 | 0.521 | .470 | .08 |
| **Intercept** | -1.086 | 0.563 | -2.189 | 0.018 | -1.929 | 0.054 | | | | |
| **Slope** | 0.001 | 0.001 | 0.001 | 0.001 | 0.722 | 0.470 | | | | |

Table S7 summarizes the results of continuous moderators. Note: SE = standard error of the coefficient; LL = lower limit of the 95% CI; UL = upper limit of the 95% CI; z = z-score associated with the g value in the same row; p = p-value associated with the z-score in the same row; k = number of effect sizes contributing to g in the same row; Q = result of the Q-test for moderation; p = p-value of the Q-test for moderation; R2 = amount of heterogeneity accounted for.

Considering the moderator (or subgroup) analysis, we could explore the effect of NiBS type (tDCS and rTMS) and the target region (lDLPFC, lM1, cpM1). In Table S4, the p values in the seventh column show whether the subgroup-specific effects are significant. Considering the NIBS type, we can see this is the case for both studies applying tDCS and rTMS. At the same time, the value under $p_{subgroup}$ shows that the difference in effects between the two subgroups is not significant. The same reasoning can be applied to the brain regions targeted by the included studies: effects were significant in the three subgroups, without statistical difference among the three. Results from the covariates are summarized in Table S5. The three covariates did not predict the studies' effect sizes.

*Emotional distress change score before and after the intervention*

As we commented in the 'Model's selection' paragraph, we selected the reduced model since it seems to better represent our data. Here, we detail results from the multi-level model, showing (i) our choice to apply the two-level model to our data and (ii) no major differences were found in the two models' results. Indeed, the multi-level model showed a small but significant effect of real stimulation in reducing emotional distress after the treatment $g$ = - 0.22, 95% CI [-0.43, - 0.01], z = - 2.08, p = .037. The meta-analysis also revealed high heterogeneity between studies $Q_{(31)} = 53.52$, p = .007, $\tau^2_{\text{between-studies heterogeneity}} = 0.12$, and $\tau^2_{\text{within-studies heterogeneity}} = 0$. Since all the variance was explained at the cluster level (between-studies heterogeneity) and no variance was explained by nesting the effect sizes within the clusters (within-studies heterogeneity), we removed this level from our analysis and applied the reduced model to our data.

**Table S8.** Subgroup analysis in pre-post emotional distress change score

| Moderator / Subgroup analysis | g | SE | LL | UL | z | p | k | psubgroup |
|---|---|---|---|---|---|---|---|---|
| **Emotional Distress pre-post** | | | | | | | | |
| **NIBS type** | | | | | | | | .799 |
| **tDCS** | -0.180 | 0.096 | -0.369 | 0.008 | -1.874 | .061 | 19 | |
| **rTMS** | -0.220 | 0.135 | -0.486 | 0.045 | -1.625 | .104 | 11 | |
| **Target region** | | | | | | | | .602 |
| **lDLPFC** | -0.296 | 0.241 | -0.769 | 0.177 | -1.227 | .220 | 6 | |
| **lM1** | -0.179 | 0.095 | -0.364 | 0.006 | -1.895 | .058 | 20 | |
| **Emotional distress symptoms** | | | | | | | | .314 |
| **Anxiety** | -0.094 | 0.143 | -0.374 | 0.187 | -0.655 | .512 | 12 | |
| **Depression** | -0.223 | 0.080 | -0.381 | -0.066 | -2.780 | .005 | 20 | |

Table S6 summarizes the categorical moderators and subgroup analyses. Note: g = Hedges' g effect size; SE = standard error of the coefficient; LL = lower limit of the 95% CI; UL = upper limit of the 95% CI; z = z-score associated with the g value in the same row; p = p-value associated with the z-score in the same row; k = the number of effect sizes contributing to g in the same row; $p_{subgroup}$ = p-value of the subgroup comparison.

**Table S9.** Covariates analysis in pre-post emotional distress change score.

| Moderator | Estimate | SE | LL | UL | z | p | k | Q | p | R2 |
|---|---|---|---|---|---|---|---|---|---|---|
| **Illness duration** | | | | | | | 19 | 3.409 | .065 | 15.66 % |
| **Intercept** | -0.494 | 0.178 | -0.843 | -0.145 | -2.773 | .006 | | | | |
| **Slope** | 0.029 | 0.016 | -0.002 | 0.061 | -1.847 | .065 | | | | |
| **Number of sessions** | | | | | | | 32 | 2.512 | .113 | 15.14 % |
| **Intercept** | -0.047 | 0.159 | -0.264 | 0.358 | 0.294 | .769 | | | | |
| **Slope** | -0.022 | 0.014 | -0.049 | 0.005 | -1.585 | .113 | | | | |

Table S7 summarizes the results of continuous moderators. Note: SE = standard error of the coefficient; LL = lower limit of the 95% CI; UL = upper limit of the 95% CI; z = z-score associated with the g value in the same row; p = p-value associated with the z-score in the same row; k = number of effect sizes contributing to g in the same row; Q = result of the Q-test for moderation; p = p-value of the Q-test for moderation; $R^2$ = amount of heterogeneity accounted for.

Considering the moderator (or subgroup) analysis, we could explore the effect of NiBS type (tDCS and rTMS), the target region (lDLPFC, lM1), and the emotional distress symptoms, measuring anxiety and depression. In Table S6, the p values in the seventh column show whether the subgroup-specific effects are significant. Considering the NIBS type, we can see that a trend existed for tDCS but not for rTMS, which included fewer studies. Considering the target region, a trend existed for lM1 but not for the lDLPFC, but even in this case, the two subgroups included different effect sizes. Considering symptoms representing emotional distress, effects were significant in the subgroup targeting depressive but not anxiety symptoms. Differences between the subgroups were significant for any of the explored moderators. Meta-regressions highlighted a trend toward significance for the illness duration. For every additional year of illness, the effect size g of a study is expected to reduce by 0.03. Therefore, we can say that the impact of real stimulation in reducing emotional distress is less effective when the year of illness increases.

*Functional disability change score before and after the intervention*

**Table S10**. Subgroup analysis in pre-post functional disability change score

| Moderator / Subgroup analysis | g | SE | LL | UL | z | p | k | psubgroup |
|---|---|---|---|---|---|---|---|---|
| **Functional Disability pre-post** | | | | | | | | |
| **NIBS type** | | | | | | | | .167 |
| **tDCS** | -0.269 | 0.075 | -0.417 | 0.122 | -3.570 | < .001 | 14 | |
| **rTMS** | -0.495 | 0.199 | -0.885 | -0.106 | -1.495 | .013 | 7 | |

Table S8 summarizes the categorical moderator and subgroup analysis. Note: g = Hedges' g effect size; SE = standard error of the coefficient; LL = lower limit of the 95% CI; UL = upper limit of the 95% CI; z = z-score associated with the g value in the same row; p = p-value associated with the z-score in the same row; k = the number of effect sizes contributing to g in the same row; $p_{subgroup}$ = p-value of the subgroup comparison.

**Table S11.** Covariates analysis in pre-post functional disability change score.

| Covariate | Estimate | SE | LL | UL | z | p | k | Q | p | R2 |
|---|---|---|---|---|---|---|---|---|---|---|
| **Illness duration** | | | | | | | 13 | 0.433 | .511 | 0% |
| **Intercept** | -0.445 | 0.186 | -0.810 | -0.080 | -2.391 | .017 | | | | |
| **Slope** | 0.012 | 0.018 | -0.024 | 0.048 | 0.658 | .511 | | | | |
| **Number of sessions** | | | | | | | 22 | 0.613 | .434 | 0% |
| **Intercept** | -0.204 | 0.155 | -0.509 | 0.100 | -1.316 | .188 | | | | |
| **Slope** | -0.010 | 0.013 | -0.035 | 0.015 | -0.783 | .434 | | | | |

Table S9 summarizes the results of continuous moderators. Note: SE = standard error of the coefficient; LL = lower limit of the 95% CI; UL = upper limit of the 95% CI; z = z-score associated with the g value in the same row; p = p-value associated with the z-score in the same row; k = number of effect sizes contributing to g in the same row; Q = result of the Q-test for moderation; p = p-value of the Q-test for moderation; $R^2$ = amount of heterogeneity accounted for.

Considering the moderator (or subgroup) analysis, we could explore only the effect of NIBS type (tDCS and rTMS). Subgroup-specific effects were significant for tDCS and rTMS studies, but no differences emerged between the two subgroups ($p = .167$). The explored covariates did not predict the studies' effect sizes (all ps > .434).

#### *Section 3: Statistical Analyses of Secondary Outcome Measures*

##### *Pain intensity at the one-month follow-up*

Thirteen effect sizes were computed. The best-fitting model was the reduced one (**Table S12**). The meta-analysis results are summarized in the forest plot (**Figure S2**). The random effects model showed an effect of real noninvasive brain stimulation on pain symptom change score *g* = - 0.57, 95% CI [-0.89, - 0.24], which is significantly different from zero, z = -3.41, p <.001. This result suggests that real stimulation may have a moderate impact on reducing the pain experienced by participants one month after the end of the treatment. The meta-analysis also revealed high heterogeneity between studies $Q_{(12)} = 36.04$, $p < .001$, $\tau^2 = 0.23$ (SE = 0.15) and $I^2 = 66.70\%$ [42.60; 91.13] (moderate to substantial heterogeneity), and PIs [-1.68, 0.54]. The Baujat plot inspection (**Figure S3**) suggested that the study of Khedr and colleagues (2017)[22] greatly contributed to the heterogeneity of the analysis. The influence analysis confirmed the study as an influential case. The effect size removal, however, only slightly reduced overall significance: g = -0.44, 95% CI [-0.71, -0.16], z = -3.13, p =.002. Concerning the publication bias, the modified Egger test showed no asymmetry b=1.67, 95% CI [-1.30, 4.63], z = - 1.50; p = .134.

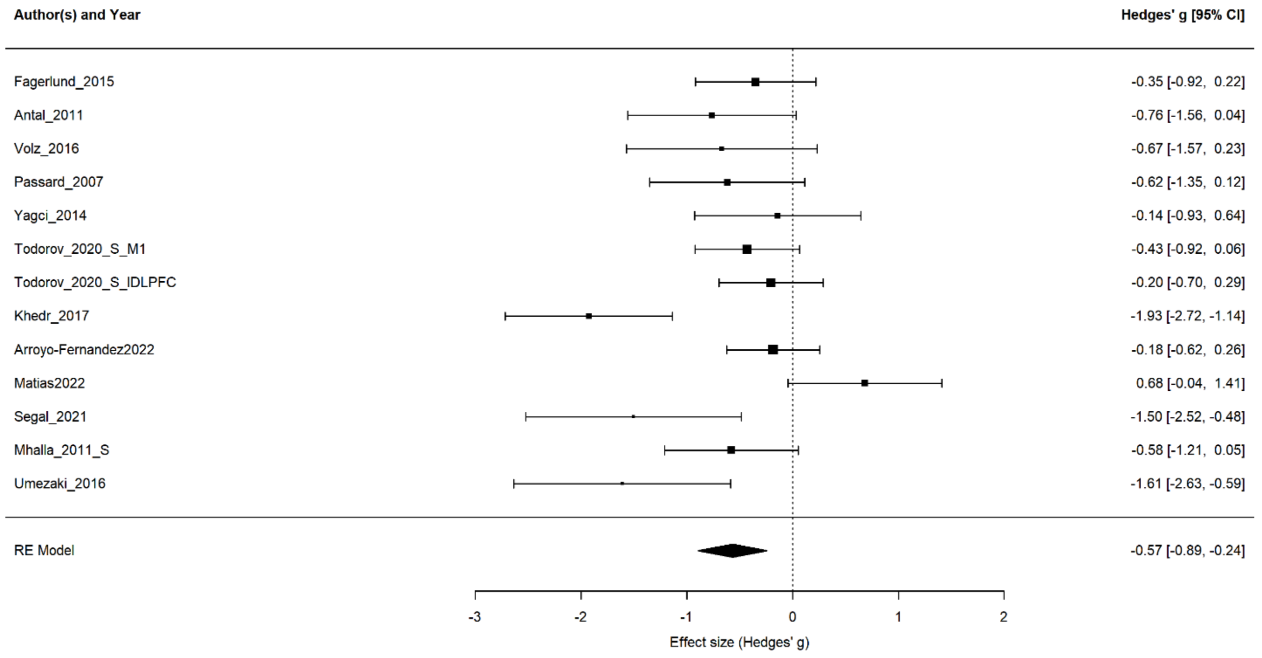

**Figure S2.** Forest plot of the effect size of noninvasive brain stimulation on pain intensity change score. CI = confidence interval.

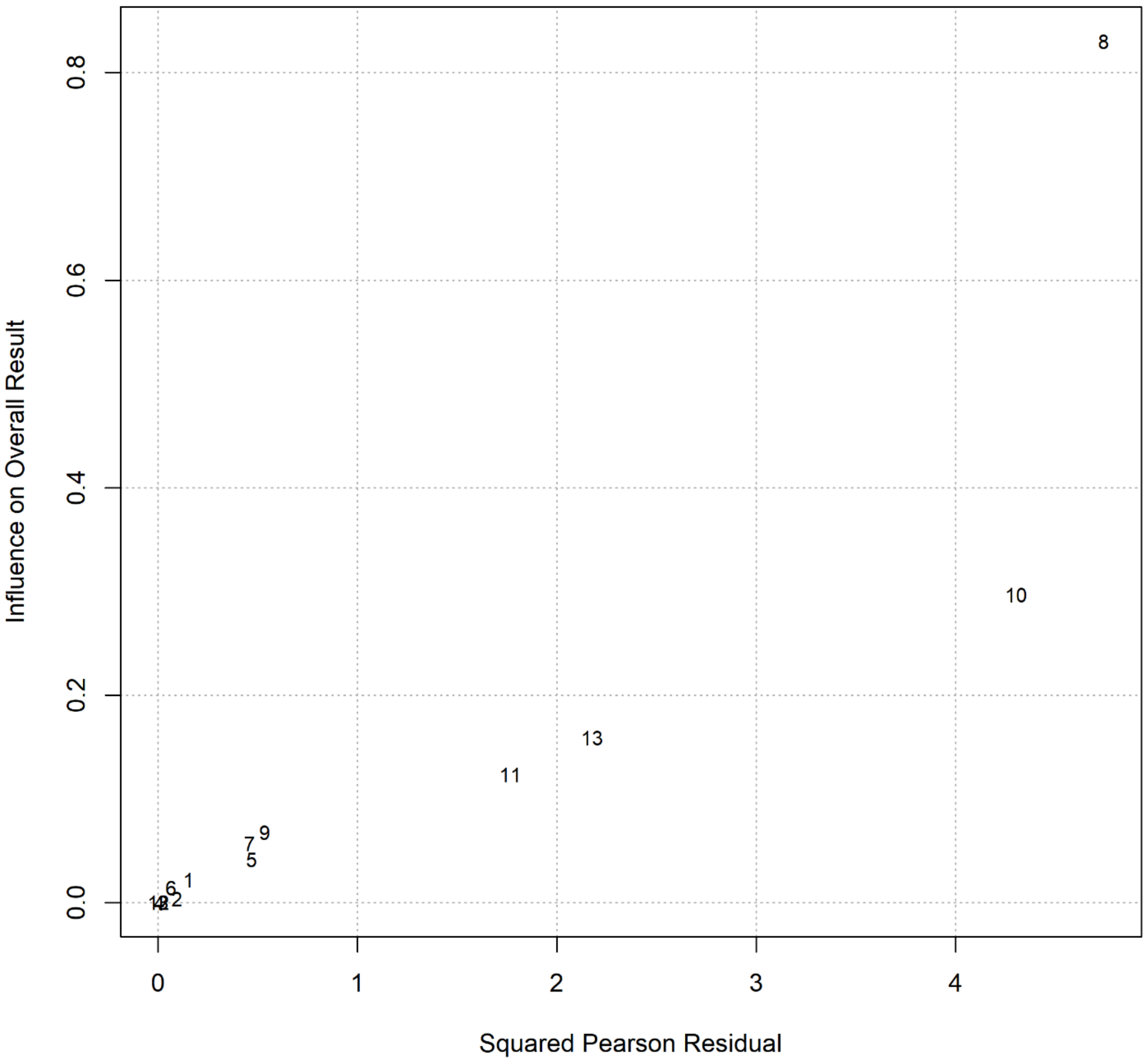


**Figure S3.** Baujat plot of studies distribution considering pain intensity at follow-up.

| **Table S12.** Subgroup analysis in pre-follow-up pain intensity change score | | | | | | | | |
|---|---|---|---|---|---|---|---|---|
| **Moderator / Subgroup analysis** | g | SE | LL | UL | z | p | k | psubgroup |
| **Pain intensity pre to follow-up** | | | | | | | | |
| **NIBS type** | | | | | | | | .851 |
| **tDCS** | -0.628 | 0.300 | -1.216 | -0.039 | -2.090 | .037 | 7 | |
| **rTMS** | -0.485 | 0.158 | -0.794 | -0.175 | -3.068 | .002 | 6 | |

Table S10 summarizes the categorical moderators and subgroup analyses. Note: g = Hedges' g effect size; SE = standard error of the coefficient; LL = lower limit of the 95% CI; UL = upper limit of the 95% CI; z = z-score associated with the g value in the same row; p = p-value associated with the z-score in the same row; k = the number of effect sizes contributing to g in the same row; $p_{subgroup}$ = p-value of the subgroup comparison.

| **Table S13**. Covariates analysis in pre-follow-up pain intensity change score. | | | | | | | | | | |
|---|---|---|---|---|---|---|---|---|---|---|
| **Moderator/coefficient** | Estimate | SE | LL | UL | z | p | k | Q | p | R2 (%) |
| **Illness duration** | | | | | | | 11 | 6.080 | .014 | 52.8 |
| **Intercept** | -1.241 | 0.302 | -1.833 | -0.649 | -4.111 | <.001 | | | | |
| **Slope** | 0.054 | 0.022 | 0.011 | 0.097 | 2.466 | n | | | | |
| **Number of sessions** | | | | | | | 13 | 3.058 | .080 | 20.31 |
| **Intercept** | 0.002 | 0.356 | -0.695 | 0.699 | 0.006 | .995 | | | | |
| **Slope** | -0.068 | 0.039 | -0.144 | 0.008 | -1.749 | .080 | | | | |

Table S11 summarizes the results of continuous moderators. Note: SE = standard error of the coefficient; LL = lower limit of the 95% CI; UL = upper limit of the 95% CI; z = z-score associated with the g value in the same row; p = p-value associated with the z-score in the same row; k = number of effect sizes contributing to g in the same row; Q = result of the Q-test for moderation; p = p-value of the Q-test for moderation; $R^2$ = amount of heterogeneity accounted for.

Considering subgroup analysis, we could explore the effect of NIBS type (tDCS and rTMS). Subgroup-specific effects were significant for tDCS and rTMS studies, but no differences emerged between the two subgroups (p = .851) (**Table S12**). The analysis of covariates (**Table S13**) highlights a significant effect of illness duration, accounting for 52.8% of heterogeneity. For every additional year of illness, the effect size g of a study is expected to reduce by 0.05. Therefore, we can say that the impact of real stimulation in reducing pain experienced at follow-up is less effective when the years of illness increase, in line with previous studies, suggesting that brain abnormalities are associated with pain duration [42] and possibly are more resistant to plastic changes induced by NIBS. Even the number of sessions showed a trend toward significance in the opposite direction. Here, for every additional session, the effect size g of a study is expected to increase by 0.07. Therefore, we can say that the impact of real stimulation in our data is augmented by increasing the number of sessions.

*Emotional distress at the one-month follow-up*

As in the primary endpoint analysis, the multi-level model was the best-fitting one, including 9 studies comprising 16 effect sizes. Even in this case, however, the analysis on the multi-level model highlighted that all the variance was due to the cluster (i.e. between study) level, $Q_{(15)} = 23.83$, p = .068, $\tau^2_{\text{Level 3}} = 0.10$ and $\tau^2_{\text{Level 2}} = 0$. Therefore, we chose the reduced model. The meta-analysis results are summarized in the forest plot (**Figure S4**). The random effects model showed no effects of real stimulation at the change score of the one-month follow-up *g* = - 0.09, 95% CI [-0.29, 0.10], z = - 0.93, p = .351. The meta-analysis also revealed low to moderate heterogeneity between studies $Q_{(15)} = 23.83$, p = .068, $\tau^2 = 0.06$ (SE = 0.06), and $I^2 = 37.06\%$ [0; 77.23], and PIs [-0.66, 0.47]. The Baujat plot inspection (**Figure S5**) suggested that the study of Khedr et al., 2017 [22] (effect size on anxiety) greatly contributed to the heterogeneity of the analysis, and the influence analysis confirmed the study as an influential case. The effect size removal, however, did not change overall results: g = - 0.02, 95% CI [-0.18, -0.14], z = -0.27 p =.788. Publication bias using the Pustejovsky-Rodgers' Egger test modification showed no asymmetry b = 0.45, 95% CI [-2.86, 3.76], which is not significantly different from zero, z = - 0.33, p = .742.

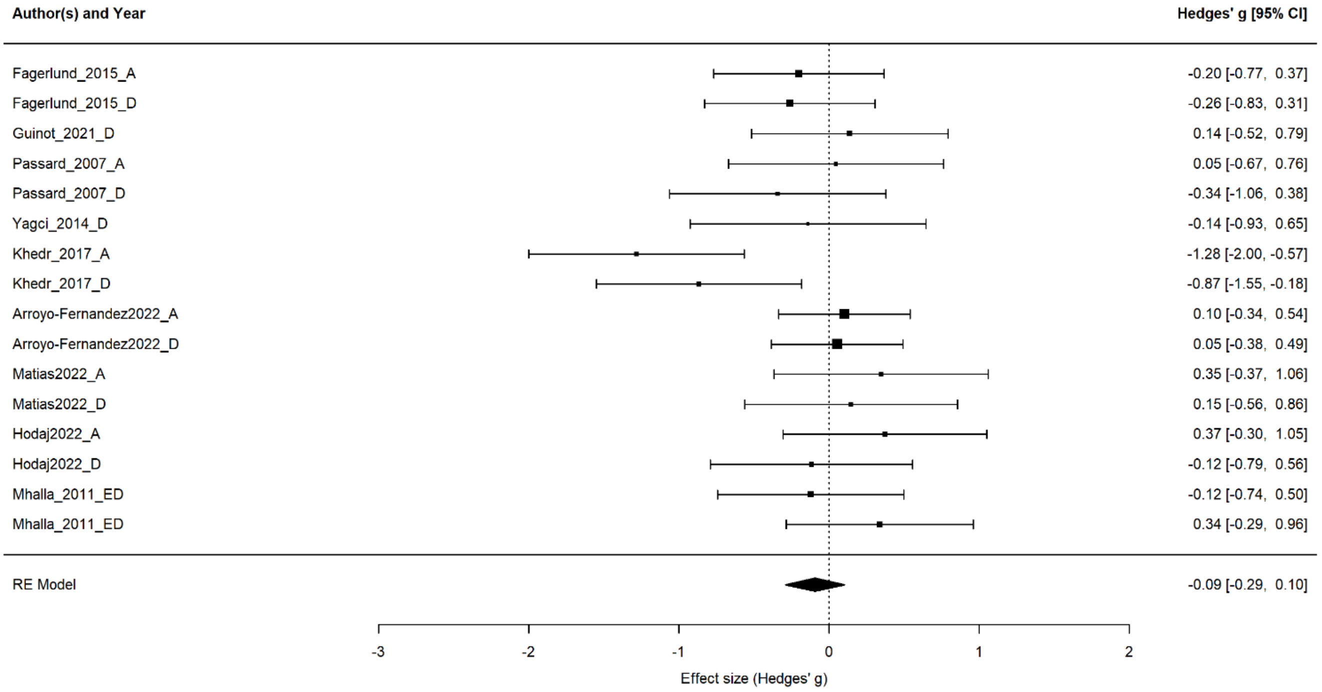


Figure S4. Forest plot of the effect size of noninvasive brain stimulation on emotional distress change score at one month of follow-up. CI = confidence interval.

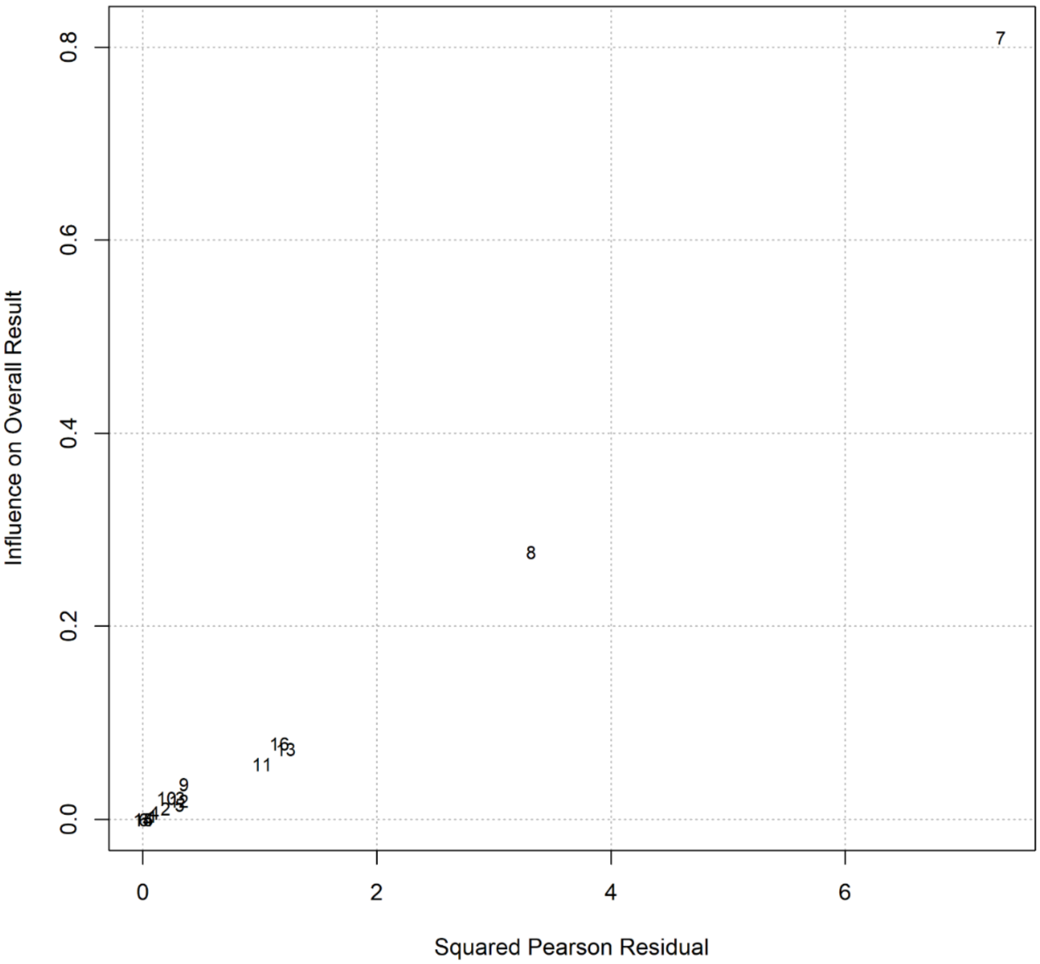


Figure S5. Baujat plot of studies distribution considering emotional distress at follow-up.

| Table S12. Subgroup analysis in pre-follow-up emotional distress change score. | | | | | | | | |
|---|---|---|---|---|---|---|---|---|
| **Moderator / Subgroup analysis** | g | SE | LL | UL | z | p | k | $p_{subgroup}$ |
| ***Emotional Distress pre-post*** | | | | | | | | |
| ***NIBS type*** | | | | | | | | .510 |
| **tDCS** | -0.152 | 0.148 | -0.441 | 0.138 | -1.027 | .305 | 10 | |
| **rTMS** | 0.006 | 0.142 | -0.271 | 0.234 | 0.045 | .964 | 6 | |
| ***Symptoms type*** | | | | | | | | .944 |
| **Anxiety** | -0.093 | 0.189 | -0.464 | 0.277 | -0.494 | .621 | 7 | |
| **Depression** | -0.093 | 0.113 | -0.314 | 0.129 | -0.817 | .414 | 9 | |

Table S12 summarizes the categorical moderators and subgroup analyses. Note: g = Hedges' g effect size; SE = standard error of the coefficient; LL = lower limit of the 95% CI; UL = upper limit of the 95% CI; z = z-score associated with the g value in the same row; p = p-value associated with the z-score in the same row; k = the number of effect sizes contributing to g in the same row; $p_{subgroup}$ = p-value of the subgroup comparison.

| Table S13. Covariates analysis in pre-follow-up emotional distress change score | | | | | | | | | | |
|---|---|---|---|---|---|---|---|---|---|---|
| **Moderator** | Estimate | SE | LL | UL | z | p | k | Q | p | $R^2$ |
| ***Illness duration*** | | | | | | | 14 | 5.911 | .015 | 62.79 % |
| **Intercept** | -0.543 | 0.195 | -0.926 | -0.160 | -2.777 | .006 | | | | |
| **Slope** | 0.033 | 0.014 | 0.006 | 0.059 | 2.431 | .015 | | | | |
| ***Number of sessions*** | | | | | | | 16 | 0.007 | .933 | 0% |
| **Intercept** | -0.075 | 0.271 | -0.605 | 0.456 | -0.276 | .783 | | | | |
| **Slope** | -0.002 | 0.028 | -0.058 | 0.053 | -0.084 | .933 | | | | |

Table S13 summarizes the results of continuous moderators. Note: SE = standard error of the coefficient; LL = lower limit of the 95% CI; UL = upper limit of the 95% CI; z = z-score associated with the g value in the same row; p = p-value associated with the z-score in the same row; k = number of effect sizes contributing to g in the same row; Q = result of the Q-test for moderation; p = p-value of the Q-test for moderation; $R^2$ = amount of heterogeneity accounted for.

Considering the subgroup analysis, we could explore the effect of NIBS type (tDCS and rTMS) and the emotional distress symptoms (anxiety and depression) (**Table S12**). No specific effects nor differences between the moderators' subgroups emerged. Covariate analysis highlighted an effect of the illness duration (p = .015) that explained the 62.8% heterogeneity **(Table S13)**. For every additional year of illness, the effect size g of a study is expected to reduce by 0.03. Therefore, we can say that the impact of real stimulation in reducing emotional distress is less effective when the year of illness increases.

*Functional disability at the one-month follow-up*

Ten effect sizes were computed. The best-fitting model was the reduced one. The meta-analysis results are summarized in the forest plot (**Figure S6**). The random effects model showed an effect of real noninvasive brain stimulation on functional disability change score *g* = - 0.51, 95% CI [-0.84, - 0.18], which is significantly different from zero, z = - 2.99, p = .003. This result suggests that real stimulation has a small to moderate impact on reducing the functional disability rated by participants at the follow-up. The meta-analysis also revealed low heterogeneity between studies $Q_{(9)}$ = 26.66, p = .002, $\tau^2$ = 0.19 (SE = 0.14), and $I^2$ = 66.24% [27.19; 90.25], and PIs [-1.58, 0.56]. Baujat plot inspection (**Figure S7**) suggested the effect size 3 (Passard et al., 2007) as a potential outlier. However, the influence analysis did not highlight influential cases.

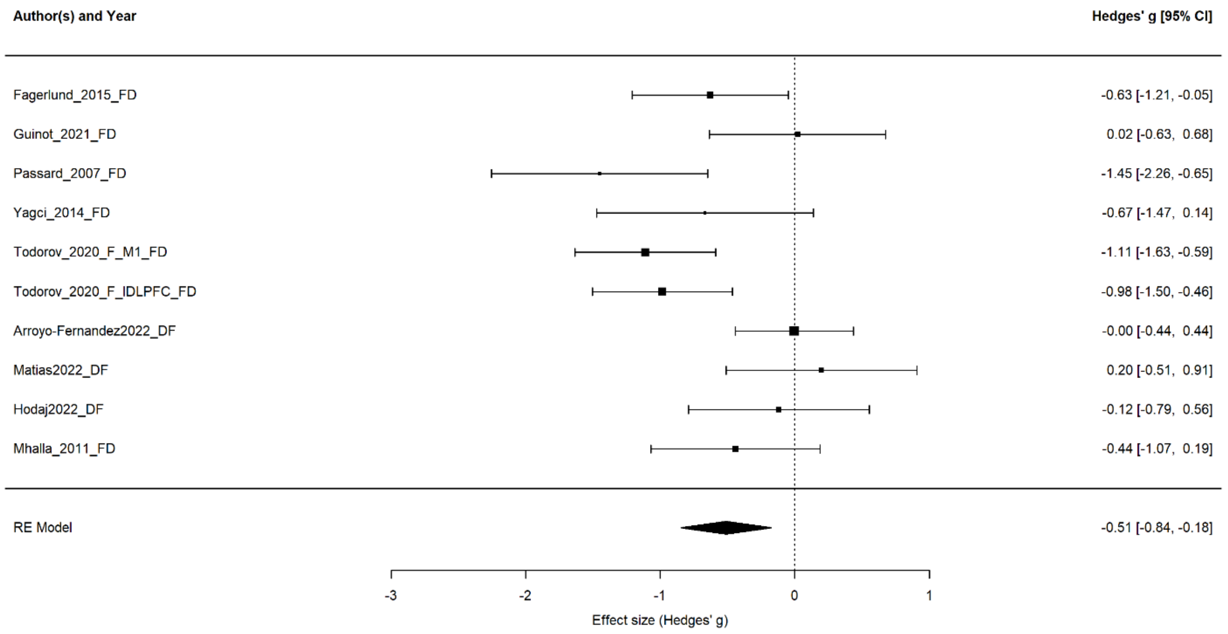


**Figure S6**. Forest plot of the effect size of noninvasive brain stimulation on functional disability change score. Negative effect sizes, in this case, represent a lower functional disability after the treatment. CI = confidence interval.

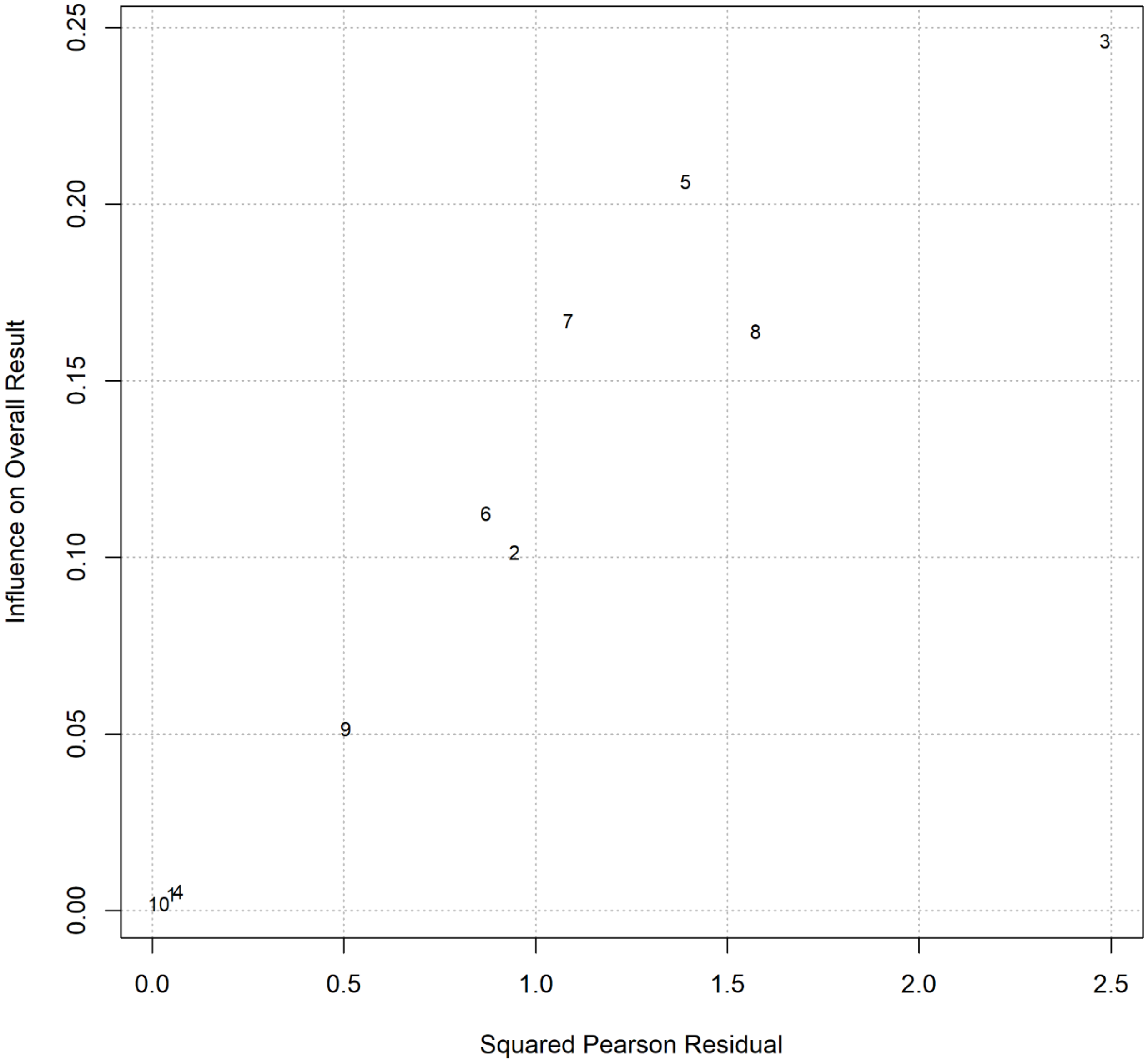


**Figure S7**. Baujat plot of studies distribution considering functional disability as the outcome measure.

Publication bias using the Pustejovsky-Rodgers' Egger test modification showed no asymmetry b = -0.92, 95% CI [-4.76, 2.93], which is not significantly different from zero, z = 0.20, p = .840. No subgroup analysis was performed due to the reduced number of studies. Covariates analysis did not highlight significant effects (Table S14).

| **Table S14.** Covariates analysis in pre-follow-up functional disability change score. | | | | | | | | | | |
|---|---|---|---|---|---|---|---|---|---|---|
| **Moderator** | Estimate | SE | LL | UL | z | p | k | Q | p | $R^2$ |
| ***Illness duration*** | | | | | | | 9 | 0.001 | .989 | 0% |
| **Intercept** | -0.576 | 0.496 | -1.547 | 0.396 | -1.162 | .245 | | | | |
| **Slope** | -0.001 | 0.031 | -0.061 | 0.060 | -0.013 | .989 | | | | |
| ***Number of sessions*** | | | | | | | 10 | 0.324 | .569 | 0% |
| **Intercept** | -0.722 | 0.411 | -1.526 | 0.083 | -1.759 | .079 | | | | |
| **Slope** | 0.025 | 0.044 | -0.061 | 0.112 | 0.570 | .569 | | | | |

Table S14 summarizes the results of continuous moderators. Note: SE = standard error of the coefficient; LL = lower limit of the 95% CI; UL = upper limit of the 95% CI; z = z-score associated with the g value in the same row; p = p-value associated with the z-score in the same row; k = number of effect sizes contributing to g in the same row; Q = result of the Q-test for moderation; p = p-value of the Q-test for moderation; $R^2$ = amount of heterogeneity accounted for.

*Quality of life change score before and after the intervention*

Seven studies included pre- and post-treatment measurements of quality of life (the paper by Fagerlund et al., 2015 included only follow-up measures; therefore, it was excluded from the analysis). The meta-analysis results are summarized in the forest plot (**Figure S8**). The random effects model did not show an effect of NIBS on the quality-of-life change score *g* = 0.30, 95% CI [-0.06, 0.67], which is not significantly different from zero, z = 1.62, p =.106. This result suggests that real stimulation does not impact on the quality of life experienced by participants after the treatment. Unlike the previous measurements, Hedge's g has a positive value since standardized questionnaires evaluating quality of life go in the opposite direction compared to scales measuring the previous symptoms, namely, higher scores represent higher quality of life. The meta-analysis also revealed moderate heterogeneity between studies $Q_{(6)} = 13.32$, p = .038, $\tau^2 = 0.13$ (SE = 0.14) and $I^2 = 54.94\%$ [0; 92.38] (moderate heterogeneity), and PIs [-0.73, 1.34]. The Baujat plot inspection (**Figure S9**) suggested that the study of Tekin et al. (2014) [40] greatly contributed to the heterogeneity of the statistical analysis. The influence analysis confirmed this study as an influential case. The effect size removal did not change the lack of impact of brain stimulation on quality-of-life scores g = 0.14, 95% CI [-0.14, 0.43], z = 0.98, p = .326. Considering the restricted number of effect sizes included in this analysis, we did not run meta-regression analyses. Moreover, since only three studies included a follow-up measure, we did not run analyses on such an effect.

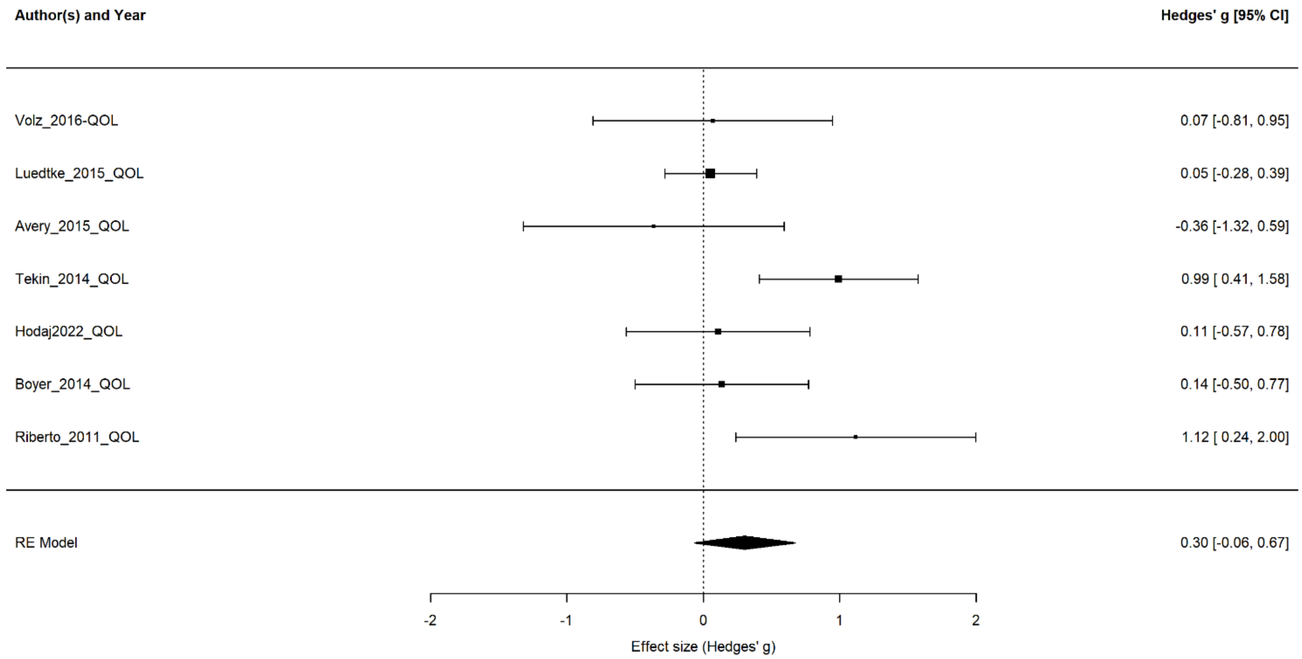


**Figure S8**. Forest plot of the effect size of noninvasive brain stimulation on quality-of-life change score. Positive effect sizes, in this case, represent a higher quality of life after treatment. CI = confidence interval.

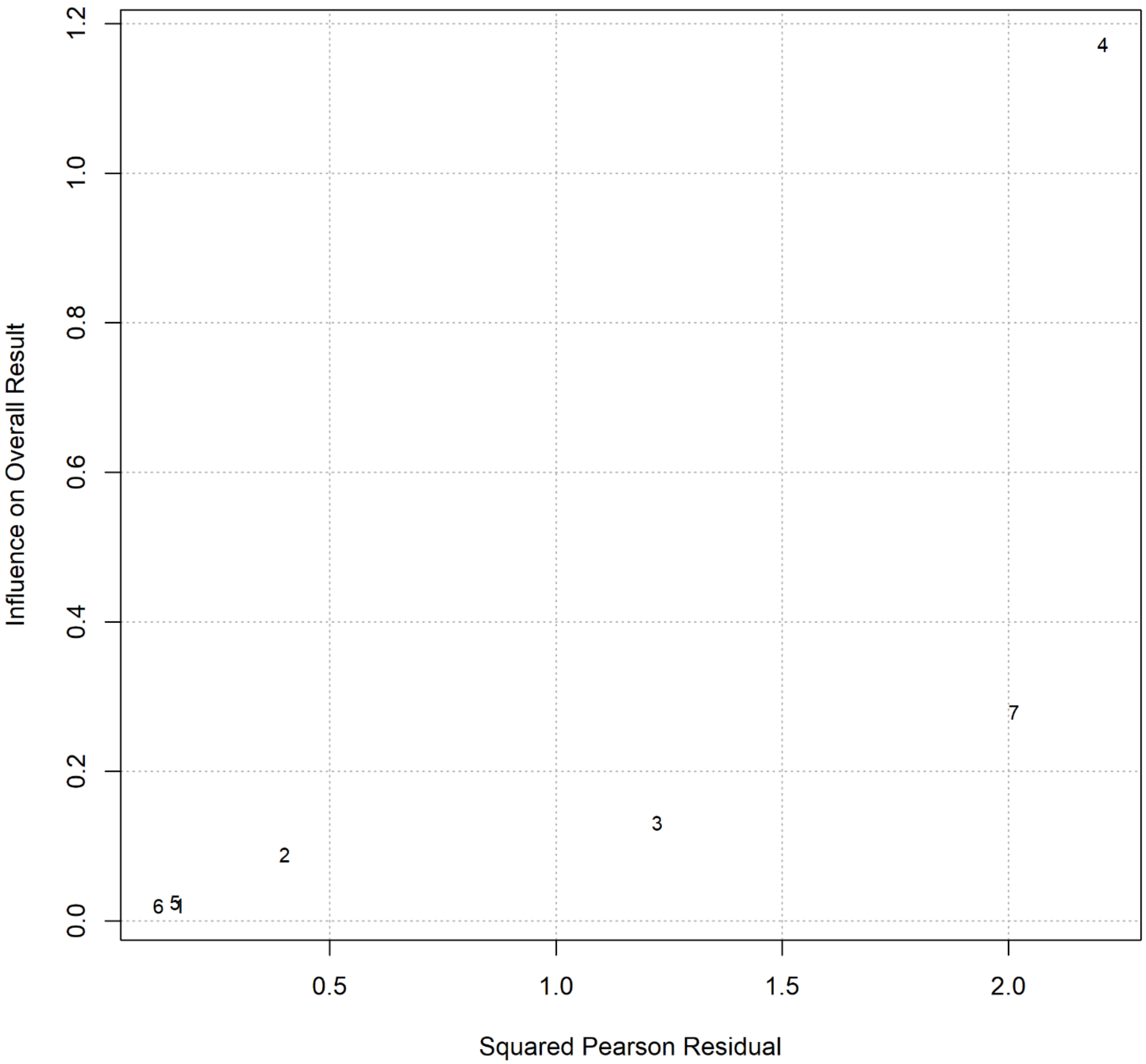


**Figure S9**. Baujat plot of studies distribution considering quality of life as the outcome measure.

*Correlations*

Correlations were performed among effect size in the three core symptoms of CPP immediately after the treatment and at follow-up, plus the effects on quality of life measured after the treatment. Considering the emotional distress, effect sizes were averaged when a study included both anxiety and depressive symptoms to produce a unique measure. Crucially, the different symptoms were differently represented numerically. Specifically, we could include 33 effect sizes for pain intensity, 13 for the pain intensity one-month follow-up, 21 for the average emotional distress, 9 for the average emotional distress at the follow-up, 22 for the functional disability, and 10 for its follow-up, 7 for the quality-of-life effect sizes. No correlation was possible between quality of life and the three core symptoms at follow-up since studies including quality of life assessment did not record the one-month follow-up included in our analyses.

Correlations highlighted a positive and significant association between pain intensity after the treatment and pain intensity and emotional distress at follow-up ($p < .001$ and $p = .001$, respectively), between emotional distress after the treatment and pain intensity and emotional distress at follow-up ($p = .027$ and $p = .001$), and between functional disability at baseline and emotional distress and functional disability at follow-up ($p = .004$ and $p = .002$). Considering follow-up measures, the effect sizes of the core symptoms correlated (ps < .050) differently from what happened immediately after the treatment (ps > .073). Quality of life displays a negative – although non-significant – pattern compared to the other symptoms since here the questionnaire scores are in the opposite direction compared to pain intensity, emotional distress, and functional disability, i.e. higher scores represent higher quality of life. Figure S10 provides a graphical representation of the correlations.

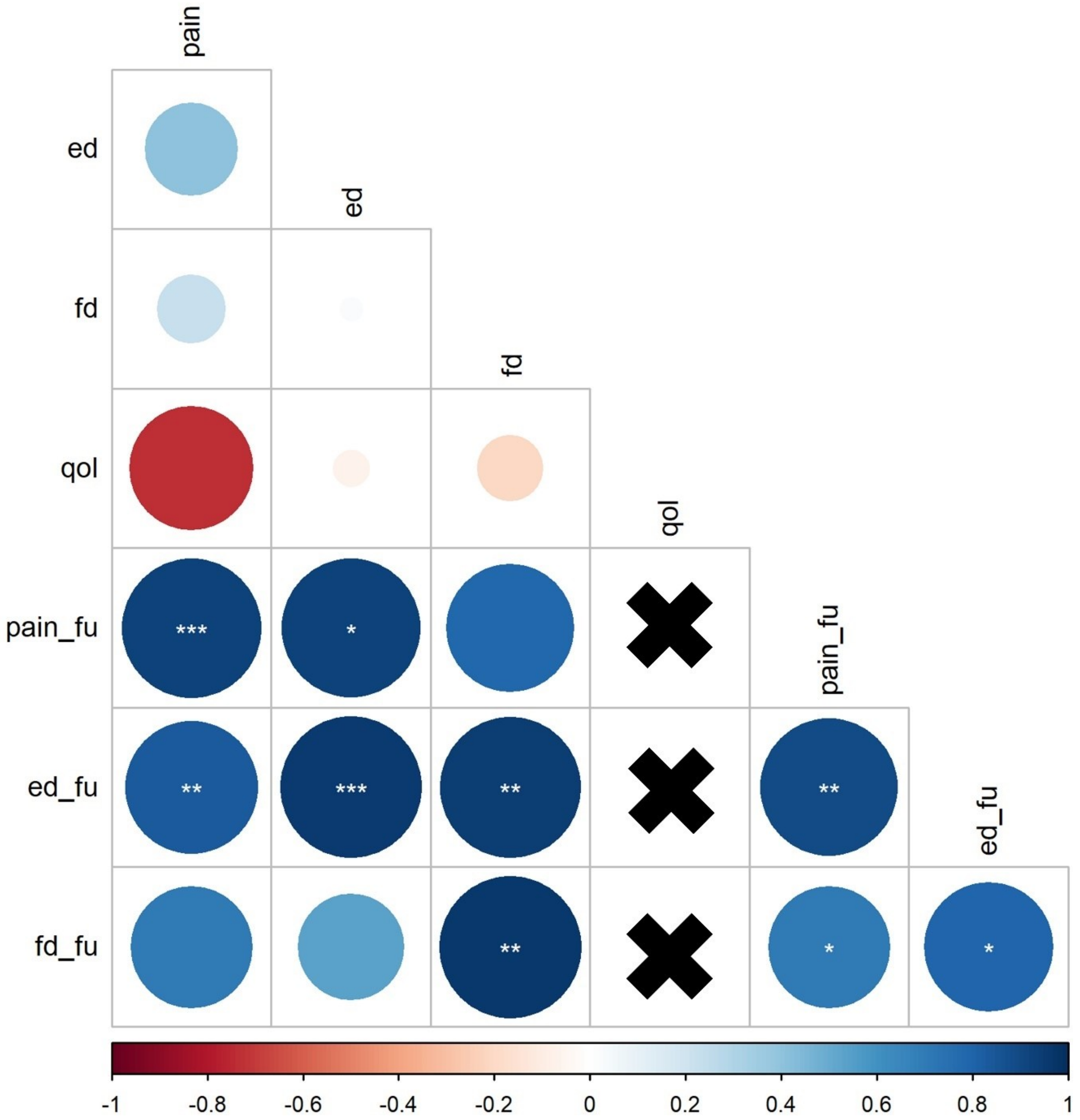


**Figure S10.** The figure shows the correlation matrix depicting the relationships between each pair of effect sizes. Positive correlations are blue-colored, and negative correlations are red-colored. Dots' color intensity and size are proportional to the correlation coefficients, and asterisks inside the dots represent the statistical significance (*$p < 0.05$, **$p < 0.01$, ***$p < 0.001$). The black cross represents the absence of data points to be correlated.

Note: ed = emotional distress; ed_fu = emotional distress at follow-up; fd = functional disability; fd_fu = functional disability at follow-up; pain = pain intensity; pain_fu = pain intensity at follow-up; qol = quality of life.
